# Exploring the Role of Plasma Lipids and Statins Interventions on Multiple Sclerosis Risk and Severity: A Mendelian Randomization Study

**DOI:** 10.1101/2022.08.01.22277781

**Authors:** Mona M. Almramhi, Chris Finan, Catherine S. Storm, Amand F. Schmidt, Demis A. Kia, Rachel Coneys, Sandesh Chopade, Aroon D. Hingorani, Nicholas W. Wood

**Affiliations:** Department of Clinical and Movement Neurosciences, University College London Queen Square Institute of Neurology, London, United Kingdom; Faculty of Applied Medical Sciences, King Abdulaziz University, Kingdom of Saudi Arabia; Institute of Cardiovascular Science, Faculty of Population Health, University College London, London, United Kingdom; British Heart Foundation University College London Research Accelerator, London, United Kingdom; Department of Cardiology, Division Heart and Lungs, University Medical Center Utrecht, Utrecht, the Netherlands; Department of Clinical and Movement Neurosciences, Queen Square Institute of Neurology, University College London, London, United Kingdom; Health Data Research UK London, University College London

## Abstract

**Background:** There has been considerable interest in statins due to their pleiotropic effects beyond their lipid-lowering properties. Many of these pleiotropic effects are predominantly ascribed to their capacity to inhibit the isoprenylation of Rho small guanosine triphosphatases (Rho GTPases). We aimed to genetically investigate the role of lipids and statin interventions on multiple sclerosis (MS) risk and severity.

**Method:** We employed two-sample Mendelian randomization (MR) to: (1) investigate the causal role of lipids (high-density lipoprotein cholesterol (HDL-C) and triglycerides (TG)) levels in MS risk and severity, (2) genetically mimic both cholesterol-dependent (via low-density lipoprotein cholesterol (LDL-C) and cholesterol biosynthesis pathway) and cholesterol-independent (via Rho GTPases) effects of statins on MS risk and MS severity.

**Results:** The results of MR using the inverse variance weighted method show that lifelong higher HDL-C (OR 1.14 (95% CI 1.04 to1.26), p-value 7.94E-03) increase MS risk, but LDL-C and TG were not. MR results also show that genetically predicted *RAC2* (OR 0.86 (95% CI 0.78 to 0.95), p-value 3.80E-03) is implicated causally in reducing MS risk. Furthermore, we found no evidence for the causal role of lipids and genetically mimicked statins on MS severity. There is no evidence of reverse causation between MS risk and lipids.

**Conclusions:** Evidence from this study suggests that HDL-C is a risk factor for MS development. The MR findings suggest that *RAC2* (a member of Rho GTPases) is a potent genetic modifier of MS risk. Since *RAC2* has been reported to mediate some of the pleiotropic effects of statins, we suggest that statins reduce MS risk via a *RAC2*-related mechanism(s) (i.e., cholesterol-independent pathway).

## Introduction

Findings from the phase 2 MS-STAT trial (a randomised, placebo-controlled trial) showed that a high dose of simvastatin (80 mg per day) led to a significant drop in brain atrophy (by 43%) and disability progression among 140 patients with secondary progressive multiple sclerosis (MS) over two years ^1^. However, whether statins’ beneficial effects on MS are mediated by cholesterol-lowering or cholesterol-independent pathway is not clear yet.

Indeed, recent evidence derived from clinical and experimental animal models of autoimmune diseases has shown that statins exert immunomodulatory and anti-inflammatory effects beyond their lipid-lowering properties that may be beneficial in autoimmune diseases such as MS ^2, 3^. Many of these effects are predominantly ascribed to statins’ capacity to inhibit the isoprenylation (also known as prenylation or lipidation) of Rho small guanosine triphosphatases (GTPases; also known as small G-proteins) ^4-6^.

Statins exert effects via Rho GTPases by two distinct mechanisms: preventing Rho proteins from localising to the membrane localisation and loading Rho proteins with GTP (**Figure 1**). By inhibiting 3-hydroxy-3-methylglutaryl coenzyme A reductase (*HMGCR*), statins prevent the synthesis of isoprenoid intermediates and the subsequent isoprenylation of Rho GTPases ^7^. This leads to the inhibition of Rho protein translocation to the plasma membrane and thus prevents the activation of their downstream effectors ^7^. The second mechanism by which statins exert effects via Rho GTPases is GTP loading, which is the conversion of Rho proteins to their active form (GTP-bound). Inhibition of isoprenoid biosynthesis by statins results in disruption of guanine nucleotide dissociation inhibitors (GDIs)–Rho GTPase binding, which provides a potential mechanism for GTP loading of the cytosolic Rho proteins ^8, 9^. GDIs are a negative regulator of Rho GTPases that only bind to isoprenylated Rho proteins to sequester them in the inactive form (GDP-bound) into the cytosol, preventing them from anchoring to membranes or being activated by guanine nucleotide exchange factors ^10^. Thus, in the absence of isoprenoid intermediates, GDIs cannot bind to Rho proteins, allowing them to be constitutively active (GTP-bound) ^10^.

**Figure 1:**
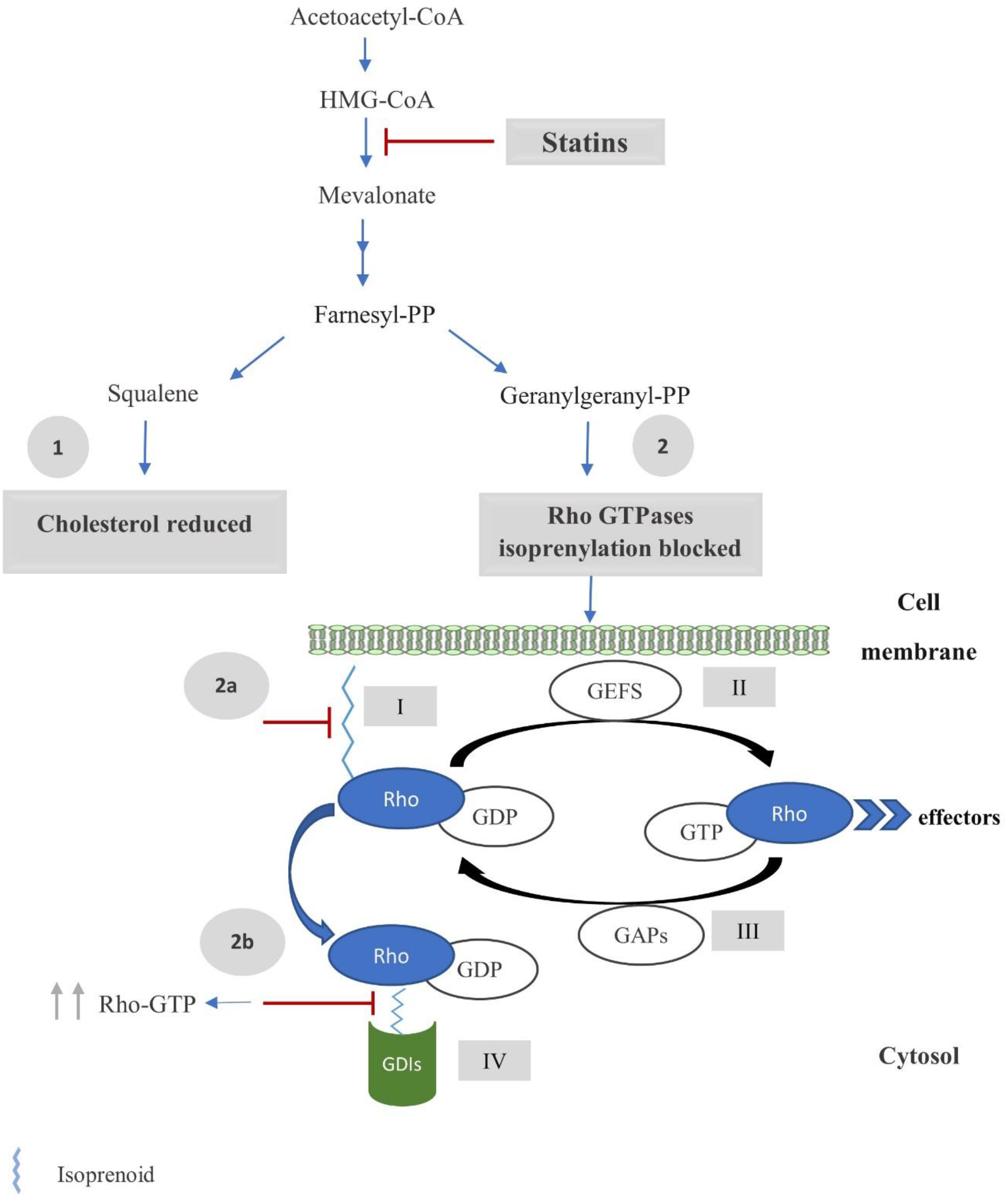
Statin effects on cholesterol and Rho GTPases. *HMGCR* inhibition by statins lead to: **(1)** reduce the synthesis of cholesterol, **(2)** and prevent the synthesis of isoprenoids (such as Farnesyl-PP and Geranylgeranyl-PP). Isoprenoids are essential molecules for the prenylation and functioning of the Rho GTPase family. After isoprenylation, the Rho proteins localise to a target cell membrane**(I)** and are activated by GEFs that facilitate the exchange of GDP for GTP **(II)**. This enables them to pass on signals to corresponding downstream effectors and regulate numerous cellular functions. Finally, the Rho proteins interact with GAPs that hydrolyse GTP to GDP, thereby inactivating the Rho proteins **(III)**. When the Rho proteins are inactivated (GDP-bound form), GDIs extract them from the membrane and sequester the proteins in the GDP-bound form into the cytosol **(IV)**. Thus, preventing the isoprenylation of Rho GTPases by statins lead to: **(2a)** the inhibition of Rho protein translocation to the plasma membrane and prevents the activation of their downstream effectors, **(2b)** and disruption of GDIs–Rho GTPase binding, which causes an increase in the levels of the cytosolic GTP-bound forms of Rho GTPases. Abbreviations: HMGCR, 3-Hydroxy-3-Methylglutaryl-CoA Reductase; Rho GTPases, Rho small guanosine triphosphatases; GEFs, guanine nucleotide exchange factors; GAPs, GTPase-activating proteins; GDIs, guanine nucleotide dissociation inhibitors.

**Figure 2:**
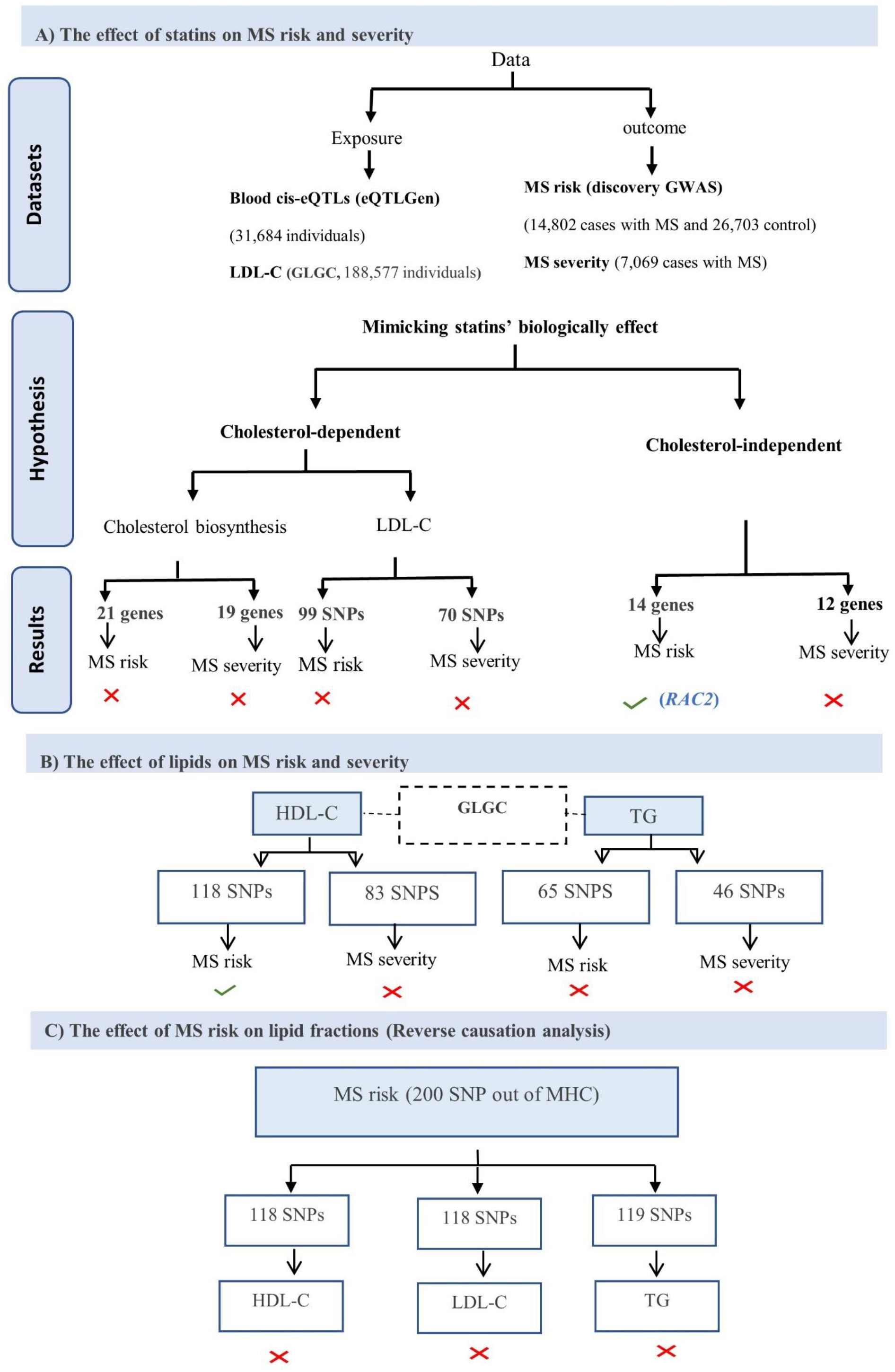
A flow diagram summarising this study’s method and results. The cross symbol indicates that there is no causal association, while the tick symbol indicates that there is a causal association. Abbreviations: GLGC, global lipids genetics consortium; MS, multiple sclerosis; HDL-C, high-density lipoprotein cholesterol; LDL-C, low-density lipoprotein cholesterol; TG, triglyceride; IMSGC, The international multiple sclerosis genetics consortium; MHC, major histocompatibility complex.

A previous Mendelian randomization (MR) analysis used single-nucleotide polymorphisms (SNPs) within *HMGCR* gene region to mimic the effects of statins on the risk of MS developing via *HMGCR* inhibition ^11^. This study revealed no causal link between these SNPs and MS risk, suggesting that statins have no effect on MS risk ^11^. *HMGCR* is the target for statins; therefore, it is not surprising that MR studies focus on *HMGCR* to mimic the effects of statins. Nevertheless, by only targeting *HMGCR*, these studies examined the cholesterol-lowering effect only and may have missed observing the statins’ pleiotropic effects. Furthermore, the effect of statins on MS severity has not yet been established. To address this knowledge gap, we adopted two-sample MR approach to genetically mimic both cholesterol-dependent and cholesterol-independent effects of statins to explore whether statins’ effects on MS risk and/or MS severity, if any, are mediated by lowering cholesterol or are independent of cholesterol. In particular, the cholesterol-dependent pathway was studied by **(a)** examining the causal role of genetically predicted the change in the blood expression levels of 25 genes (including the *HMGCR* gene) that encode proteins involved in cholesterol biosynthesis, and **(b)** examining the causal role of genetically predicted LDL-cholesterol, given that LDL-C is a relevant prognostic factor for assessing the degree of *HMGCR* inhibition. The cholesterol-independent pathway was studied by examining the causal role of genetically predicted the change in the blood expression levels of 20 genes that encode Rho GTPase family members. We sought also to examine the causal role of genetic predisposition to increased other major plasma lipid fractions (high-density lipoprotein cholesterol (HDL-C) and triglycerides (TG)) in MS risk and severity. In addition, the reverse causation between HDL-C, LDL-C, TG and MS risk is addressed in this study. Since no single loci achieved genome-wide significance in MS severity data, we were unable to perform a reverse causation between lipid fractions and MS severity.

We tested 2 hypotheses to examine whether statins influence MS through cholesterol-dependent or cholesterol-independent pathways:

1. We would expect statins causally influence MS via lowering blood cholesterol levels if we obtain:
  a. A statistically significant causal estimates for MR analyses involving LDL-associated SNPs.
  b. A statistically significant causal estimates for MR analyses involving SNPs of HMGCR and any other downstream genes involved in cholesterol biosynthesis.
2. In contrast, we would expect statins causally influence MS via cholesterol-independent pathway, if we obtain a statistically significant causal estimates for MR analyses involving SNPs of Rho GTPases.

In simple terms, MR is a type of “instrumental variable” analysis that uses genetic variants, such as SNPs, robustly associated with exposures as proxies for the risk factors of interest to investigate their causal effect roles on outcomes ^12^. MR is a useful method to appraise causality within observational epidemiology, which is relatively quicker and easier than randomized controlled trial studies and overcomes some of the limitations inherent in conventional epidemiologic studies ^13^.

## Method

### Genetic instruments selection for exposures

The summary statistics data for SNPs associated with blood lipid fractions at p-values < 5 × 10^−8^ were taken from Global Lipids Genetics Consortium genome-wide association study (GWAS) to investigate the association between lipids and MS ^14^.

To explore the reverse causation between lipid fractions and MS risk, we initially selected 200 autosomal susceptibility SNPs outside the major histocompatibility complex (MHC) region that reported by the International Multiple Sclerosis Genetics Consortium (IMSGC) as genome-wide significant for MS ^15^. With MS risk -associated SNPs as the exposure, we obtained corresponding effect estimates for HDL-C, LDL-C and TG from GLGC as the outcome.

All the selected SNPs for lipid fractions and MS risk (as exposure) were clumped at a linkage disequilibrium (LD) threshold value of *r*^*2*^ < 0.01. Then, we used Steiger filtering to remove genetic variants that explained more of the variation in the outcome than the variation in the exposure of interest ^16, 17^.

The remaining SNPs were used to calculated the mean F-statistic and the proportion of variance explained (*R*^2^) to evaluate the strength of the selected variants ^18^. The value of the mean F-statistics more than 10, indicates that bias due to weak instruments is negligible ^18^.

To investigate the potential role of and mechanisms used by statins in MS risk and severity, expression quantitative trait loci (eQTL) data with p-values < 5 × 10^−8^ were obtained from the eQTLGen to genetically mimic statin effects ^19^. We used whole-blood *cis*-eQTL in a ±5 kilobases flank around 25 genes (including *HMGCR*) that encode proteins involved in cholesterol biosynthesis and around 20 Rho GTPase gene regions to genetically mimic the effects of statins elicit via the cholesterol-dependent and cholesterol-independent pathways, respectively, (Supplementary **Table S1**). All the selected SNPs clumped at the liberal LD-clumping threshold value of *r*^*2*^ < 0.4.

For replication purpose, we obtained independent summary statistics data for lipid fractions from the Neale Lab consortium and for eQTL data from the Genotype-Tissue Expression (GTEx) project (version 8) ^20^.

### Genetic instruments selection for outcome

The summary statistics data from the discovery cohorts of the most recent MS risk GWAS were obtained from the IMSGC ^15^. Due to complex LD structures and a high potential for pleiotropy in the MHC region, 12 Mbps around this region (from 24 to 35 megabase pairs of chromosome 6; GRCh37) were excluded from MS discovery GWAS. For MS severity, we obtained the summary statistics data from the corresponding author of the original publication ^21^ (Supplementary).

### MR analysis

To assess a potential effect of the exposure of interest on the outcome, we first used the inversevariance weighted (IVW) method which in the absence of directional pleiotropy, it provides a robust causal estimates ^22^. Then, we used the MR–Egger approach, as a sensitivity analysis to detect the possible pleiotropy effects and to account for it ^22^. Because many of the SNPs were associated with more than one lipid fraction, multivariable MR (MVMR) through IVW was used to accounts for the potential pleiotropic influence ^23^. For *cis*-eQTL data, where the genetic variants are in a moderate LD (*r*^*2*^ < 0.4), we implemented the IVW and MR-Egger methods suggested by Burgess et al., which account for a correlation structure between genetic variants, thus avoiding ‘double counting’ of variant effects ^24^.

To assess the heterogeneity, we used Cochran’s Q statistic and the related *I*^*2*^ index to facilitate heterogeneity interpretation that expresses the amount of heterogeneity as a percentage ^25^. The MR-Egger intercept was used to assess the presence of pleiotropic effects, a statistically significant intercept term (p-values < 0.05) indicates directional pleiotropy ^25^.

Correcting for multiple testing was performed on IVW results using the Benjamini–Hochberg method to identify significant associations (false discovery rate (FDR) ≤ 0.05) ^26^. Results with FDR ≤ 0.05 considered having strong evidence.

## Results

### Genetically predicted HDL-C associated with increased MS risk but not severity

**Table 1** presents the number of SNPs, the explained variance (*R*^*2*^) and the mean F-statistics for each lipid trait. MR analysis was performed for each of the lipid fractions in turn, and the results of these analyses are displayed in **Figure 3A** and **4** and in supplementary **Table S2** and **Figure S1**. For HDL-C, assessment through IVW showed evidence that raised HDL-C is associated with an increase in MS risk. The MR-Egger analysis results replicated this finding. The heterogeneity was significant (Cochran’s Q p-value < 0.05). However, since the MR-Egger intercept indicates a balanced horizontal pleiotropy (p-value > 0.05), this heterogeneity is not due to pleiotropic variants. Instead, it is possibly due to a different SNP–HDL-C influence on MS risk mediated via a different biological mechanism. The MVMR analysis results after adjustment for LDL-C and TG remained broadly consistent with the primary findings in the IVW estimator, which further supported the causality relationship between HDL-C and MS risk. For LDL-C and TG, there was no evidence for a causal relationship with MS risk found in the IVW, MR-Egger and MVMR estimator results. There was evidence of heterogeneity; however, the MR-Egger intercept test did not provide any evidence of horizontal pleiotropy in these results.

**Table 1:** Sample characteristics of the lipid traits

| Lipid trait | Lipid-MS risk |  |  | Lipid-MS severity |  |  |
| --- | --- | --- | --- | --- | --- | --- |
| | No. of SNPs | $R^2$ (%) | Mean F-statistics | No. of SNPs | $R^2$ (%) | Mean F-statistics |
| **HDL-C** | 118 | 9 | 124 | 83 | 6.9 | 139 |
| **LDL-C** | 99 | 11.6 | 159 | 70 | 7.7 | 156 |
| **TG** | 65 | 6.3 | 159 | 46 | 5.2 | 189 |

HDL-C, high-density lipoprotein cholesterol; LDL-C, low-density lipoprotein cholesterol; TG, triglyceride; No. of SNPs, the number of independent genome-wide significant single nucleotide polymorphisms; $R^2$ (%), approximate variance explained by SNPs in the target trait that expressed in percentage.

**Figure 3:**
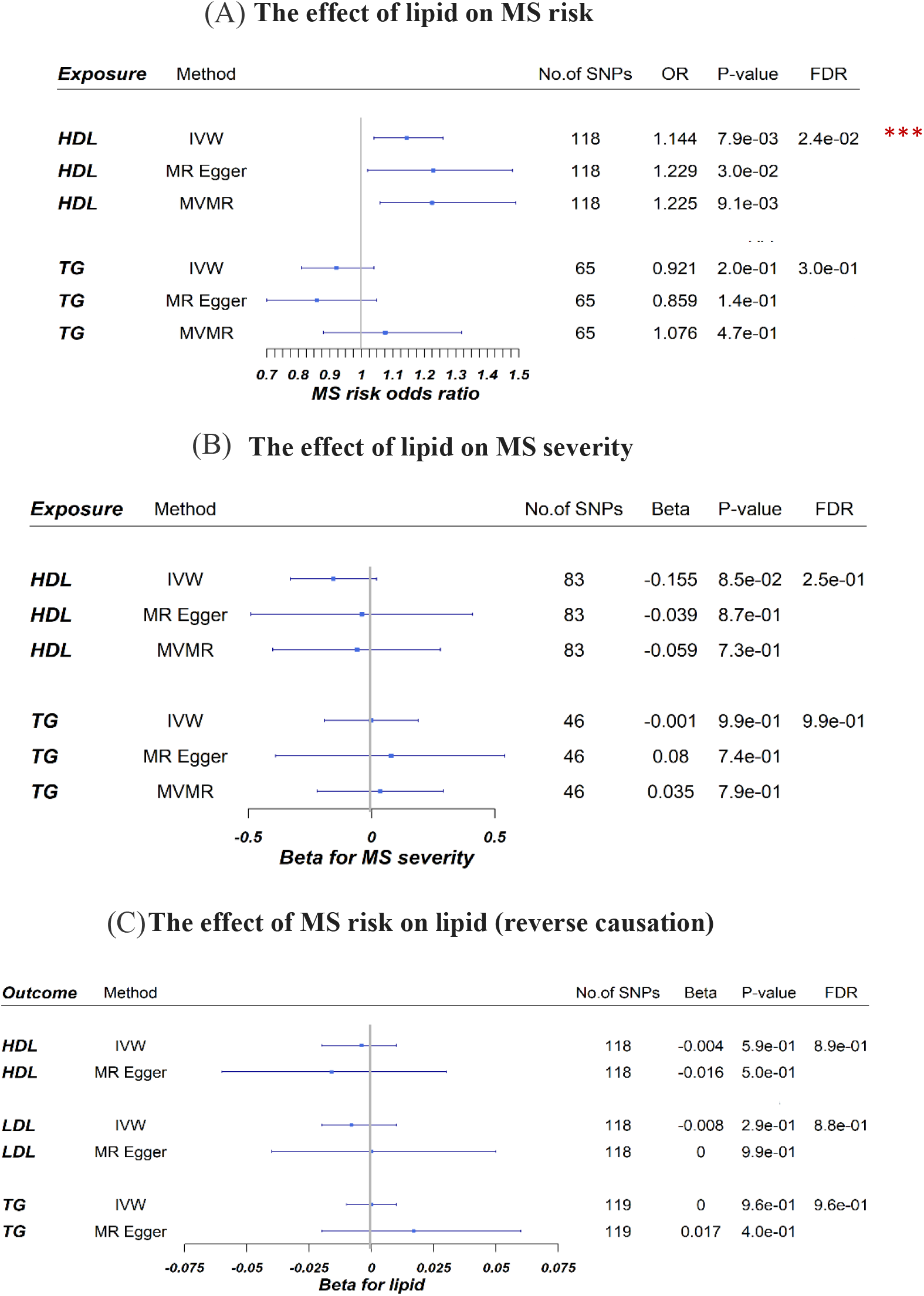
**A** forest plot showing the associations between genetically predicted lipid fractions and MS risk that reported as odds ratio per one standard deviation (SD) increase of lipid fraction. **B** forest plot showing the associations between genetically predicted lipid fractions and MS severity that reported as beta per 1-SD increase of lipid fraction. **C** forest plot showing the associations between genetically predicted MS risk and the lipid fractions which presented as beta per 1-unit-higher log-odds of MS risk. The horizontal line represents a 95% confidence interval error bars. Abbreviations: IVW, Inverse variance weighted; MVMR, Multivariable Mendelian randomization; OR, odds ratio; FDR, false discovery rate; No. of SNPs, the number of genome-wide significant single nucleotide polymorphisms.

**Figure 4:**
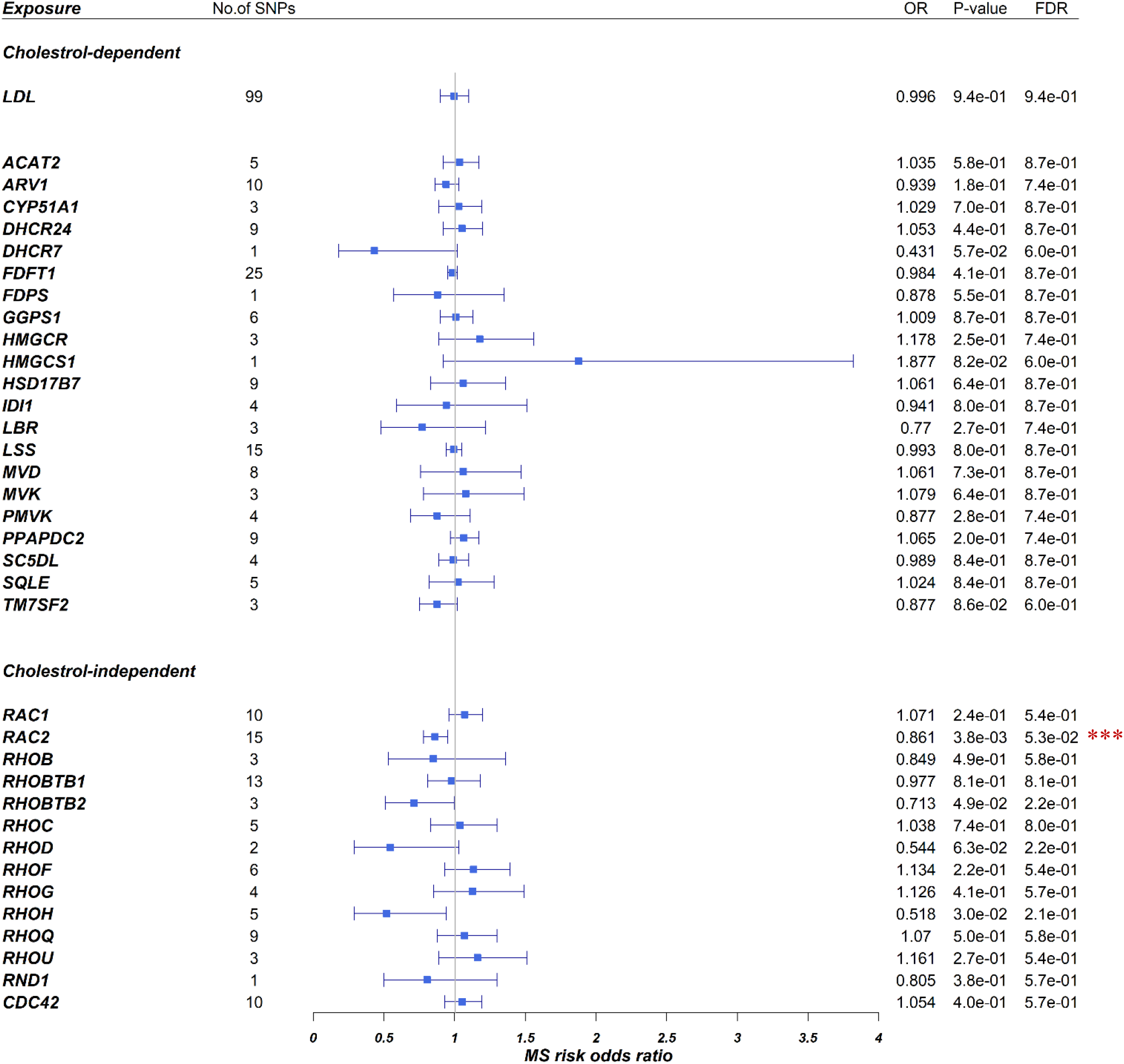
Forest plot showing the associations between the genetically mimicked statins’ biological effects via cholesterol-dependent (through LDL-C and cholesterol biosynthesis pathway) and cholesterol-independent (through Rho GTPases) and MS risk. Results from the Wald ratio (if the number of SNPs < 2) or IVW are shown. Each point represents MS’s disease odds ratio per one standard deviation increase in LDL-C level or gene expression in blood with a 95% confidence interval error bars. Abbreviations: LDL-C, low-density lipoprotein cholesterol; OR, odds ratio; FDR, false discovery rate; No. of SNPs, the number of genome-wide significant single nucleotide polymorphisms.

Since the HDL-C results were deemed significant (FDR ≤ 0.05) after multiple testing corrections, the results were assessed for replication using independent HDL-C data from UK Biobank. The replication result aligned with the initial results, further supporting the significant causal association between HDL-C and MS risk (**Table 2**). However, we must acknowledge that there was unbalanced pleiotropy in this analysis. Thus, in this case, the MR-Egger estimate, which is robust to pleiotropic instruments, is more reliable than the IVW estimate.

**Table 2:** Replication analysis results for the effect of HDL-C and *RAC2* on MS risk

| Trait | Method | No. of SNP | OR (95 % CI) | p-value | Q p-value | $I^2$ (%) | MR-Egger intercept | MR-Egger intercept p-value |
| --- | --- | --- | --- | --- | --- | --- | --- | --- |
| HDL-C | IVW | 482 | 1.08 (0.99,1.17) | 6.63E-02 | 6.43E-04 | 18.1 |  |  |
|  | MR Egger | 482 | 1.19 (1.05,1.36) | 7.23E-03 | 9.17E-04 | 17.6 | -0.0034 | 0.0484 |
| *RAC2* | IVW | 2 | 0.7 (0.51,0.96) | 2.80E-02 |  |  |  |  |

HDL-C, high-density lipoprotein cholesterol; No. of SNPs, the number of independent genome-wide significant single nucleotide polymorphisms; IVW, inverse-variance weighted; OR, odds ratio; CI, confidence interval; Q p-value, Cochran's Q statistic; FDR, false discovery rate; $I^2$ (%) expresses the level of heterogeneity as a percentage.

The IVW, MR-Egger and MVMR methods were also implemented to assess the lipid influence on MS severity. The results revealed no evidence of HDL-C, LDL-C or TG having a causal role in MS severity (**Figure 3B** and **5** and supplementary **Table S3** and **Figure S2**. No evidence of heterogeneity or pleiotropy was detected in this analysis.

### Genetically predicted MS risk not associated with lipid levels (reverse causation analysis)

We sought to explore whether the liability to MS risk would exert a change in lipid levels. To do so, we selected 118 and 119 SNPs out of the 200 that account for almost 19% of the MS heritability. The mean F-statistics of these SNPs was around 75. The IVW and MR-Egger results revealed no causal link between the genetic determinants of MS risk and HDL-C, LDL-C or TG (**Figure 3C** and supplementary **Table S4** and **Figure S3**). There was evidence of significant heterogeneity; however, the MR-Egger intercept test suggested no evidence of pleiotropy.

### Genetically mimicked effect of statins on MS risk is independent of cholesterol pathway

Figure 4 and supplementary **Table S5** and **Figure S4** display the associations between the genetically mimicked statin effects and MS risk. A total of 35 genes (21/25 genes of the cholesterol biosynthesis pathway and 14/20 genes of the Rho GTPase family; supplementary **Table S1**) were selected for analysis on the basis of having at least one SNP strongly associated with their expression. MR analyses involving SNPs in these gene regions found only a link between the expression levels of *RAC2* and MS risk that was significant (FDR ≤ 0.05) after multiple testing corrections.

The heterogeneity, in general, in these analyses ranged from non-significant to moderate, and the MR-Egger intercept test provided no evidence for horizontal pleiotropy except for *RHOH*.

For *RAC2*, the IVW result revealed that one SD genetically predicted that expression of *RAC2* in the blood was associated with a 14% reduction in MS risk. The MR-Egger causal estimate was significant and largely consistent with the IVW results, reducing the probability that pleiotropy influenced these results. There was no evidence for heterogeneity, and the MR-Egger intercept test provided no evidence for directional pleiotropy. Since the results were deemed significant after multiple testing corrections, replication was assessed using the whole-blood *cis*-eQTL dataset from the GTEx project. It was found that the direction of the effect was identical across the discovery and replication results, providing further support for *RAC2* playing a protective role in MS risk (**Table 2**).

### Genetically mimicked effect of statins had no causal association with MS severity

Figure 5 and supplementary **Table S6** display the associations between the genetically mimicked statin effects and MS severity. A total of 31 genes (19/25 genes involved in the cholesterol biosynthesis pathway and 12/20 genes of the Rho GTPase family; supplementary **Table S1**) were selected for analysis on the basis of having at least one SNP strongly associated with their expression. The MR results showed no evidence of an association between the SNPs in these genes and MS severity. There was no evidence for heterogeneity or horizontal pleiotropy in these MR analyses.

**Figure 5:**
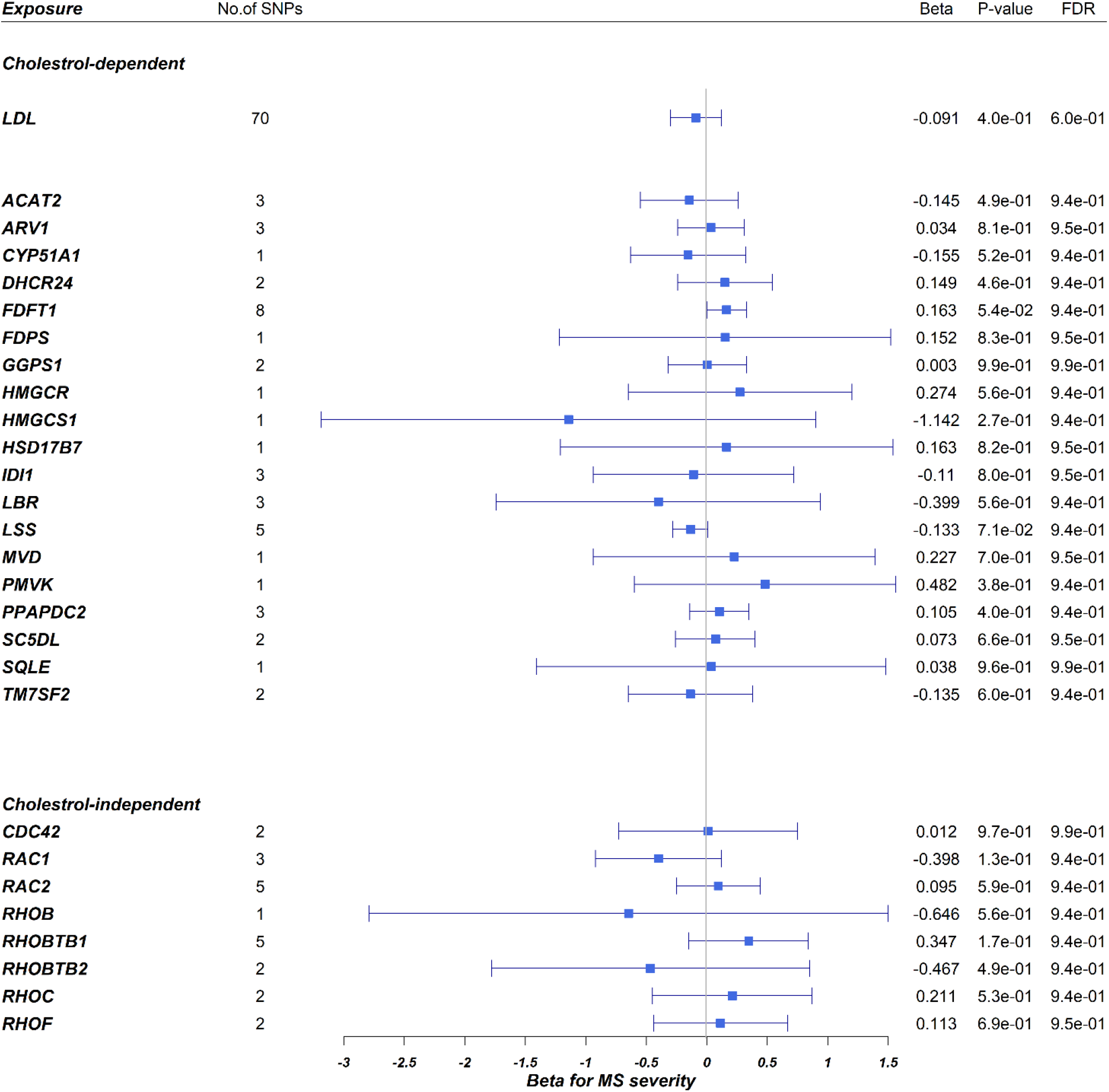
Forest plot showing the associations between the genetically mimicked statins’ biological effects via cholesterol-dependent (through LDL-C and cholesterol biosynthesis pathway) and cholesterol-independent (through Rho GTPases) and MS severity. Results from the Wald ratio (if the number of SNPs < 2) or IVW are shown. Each point represents MS’s disease log odds ratio per one standard deviation increase in LDL level or gene expression in blood with a 95% confidence interval. Abbreviations: LDL-C, low-density lipoprotein cholesterol; OR, odds ratio; FDR, false discovery rate; No. of SNPs, the number of genome-wide significant single nucleotide polymorphisms.

## Discussion

The work presented in this study aimed to: (1) dissect the causal nature of the association between blood lipid levels and MS (risk and severity) and explore whether genetic predisposition to increased major plasma lipid fractions plays an aetiological role in MS; (2) explore the potential effects of statins on MS via MR analysis conducted using SNPs in different gene regions that genetically mimic statin biological effects; and (3) assess whether there is reverse causation between lipid fractions and MS risk.

### High plasma HDL-C is a risk factor for MS

The results show that lifelong high HDL-C leads to an increased MS risk. This finding is reproducible and robust in terms of heterogeneity, pleiotropy and reverse causation testing. In contrast, genetically raised circulating TGs are unlikely to be associated with the risk of developing MS.

Associations between lipids and MS risk have received insufficient attention in epidemiological studies. Surprisingly, only one MR analysis on lipids and MS risk with GLGC and IMSGC data, the same datasets used in the current study, has been published ^27^. The primary findings of that study demonstrated that there is no causal role for genetically raised LDL-C and TGs on MS risk, and there was only weak evidence of association between genetically raised HDL-C and MS risk (IVW OR = 1.14, p-value = 0.057) ^27^.

The MR results of the current study agree with the above study regarding LDL-C and TGs but not HDL-C—we found robust evidence of an HDL-C–MS risk association. The most notable difference is the number of SNPs included in the analysis model, which may explain why previous results differ from current results regarding HDL-C. In the aforementioned study, 68 SNPs were used to genetically proxy circulating levels of HDL-C, and they explained about 1.6% of the variance in HDL-C levels. In the current study, we used 118 SNPs to genetically proxy circulating levels of HDL-C, and they explained about 9% of the variance in HDL-C levels, clearly more than the variances explained by the 68 SNPs in the previous MR study. Thus, the MR model used here had sufficient power to detect a causal association between HDL-C and MS risk.

### The lower cholesterol levels induced by statins have no effect on MS risk

In the second part of this work, we conducted a separate MR analysis to address the causal link between genetically mimicked statin effects and MS. First, we used variants in *HMGCR* and other downstream genes to mimic the cholesterol-dependent effects of statins in relation to MS risk. The findings suggest that stains have no effect on MS risk through mechanisms that contribute to cholesterol level reduction. This result was expected, because LDL-C itself does not have a causal role in MS risk in the current results and therefore using a drug intended to lower cholesterol as a therapeutic strategy will be an ineffective approach for MS prevention. Indeed, a recent study suggests that the beneficial effects of simvastatin in patients with MS are independent of serum cholesterol ^28^. In that study, the authors reanalysed the phase 2 MS-STAT trial by applying structural equation models to examine whether the beneficial effects of simvastatin on reducing the rate of brain atrophy and slowing deterioration are dependent on or independent of blood cholesterol reduction ^28^.

### The effects induced by statins via the cholesterol-independent pathway (*RAC2*) may reduce MS risk

Since the cholesterol-dependent pathway showed no effect on MS risk, our attention was directed to exploring the causal link between Rho GTPases (i.e., mimicking the independent-cholesterol effect of statins) and MS risk. Interestingly, the MR results showed that genetically predicted *RAC2* expression was causally associated with reducing MS risk, and this finding emerged as robust with sensitivity analysis and was replicated in an independent eQTL dataset (GTEx).

*RAC2* is a Rho GTPase family member (supplementary **Table S1**) expressed mainly in blood cell lineages^29^. *RAC2* regulates multiple key processes of inflammatory responses, including dendritic cell migration, nicotinamide adenine dinucleotide phosphatase oxidase activity and T-cell proliferation, migration and differentiation to the Th1 subtype ^30, 31^. In addition to immune activation, *RAC2* is involved in the induction of peripheral immune tolerance. It is an essential component of restimulation-induced cell death ^32^, a necessary process in the self-limiting negative feedback mechanism used to control T-cell expansion during ongoing immune responses ^33^.

The exact mechanisms underlying the protective role for *RAC2* in MS risk has not yet been elucidated; however, an association between *RAC2* and MS has previously been reported ^31, 34^. For example, the expression level of *RAC2* in whole blood samples from patients with MS were found to be low compared to those in healthy controls ^34^. This finding supports the protective role of *RAC2* on MS risk that we observed in the current results.

Recent findings suggest that the *RAC2* represents a pleiotropic effect of statin therapy. It has been shown that statins, through inhibition of isoprenylation of Rac2, reduce oxidative stress during sepsis and downregulate pentraxin 3 in vascular cells during immune-inflammatory responses ^35-37^. Furthermore, statins have been shown to induce the expression of several genes, including *RAC2*, that are involved in epidermal growth factor signalling ^38^; however, the mechanism by which statins can induce *RAC2* expression remains to be identified.

Taken together, the current results shed light on the role *RAC2* plays as a potent genetic modifier of MS risk. In addition, suggest that statins might mediate some beneficial effects on MS risk via *RAC2*-regulated pathways. Nonetheless, caution should be taken to avoid overinterpretation of these findings. Although MR is a powerful tool for investigating the causal relationship between an exposure and an outcome, this approach cannot identify the specific molecular mechanism(s) of the relationship or confirm the hypothesis in the current study regarding statins, *RAC2* and MS risk. In addition, the possibility that *RAC2* reducing the risk of MS is independent of statins effect cannot be ruled out. Thus, further studies are required to identify the mechanism responsible for the observed causal relationship between *RAC2* and MS risk and to test the hypothesis that statins reduce MS risk via a RAC2-related mechanism.

### No causal role for genetically mimicked effects of statins and plasma lipids in MS severity

Despite several epidemiological studies investigating the associations between circulating lipid fractions and accrual of disability in patients with MS, most of these studies used expanded disability status scale (EDSS) to measure the disability and a few used MS severity scores. The difference between these measures is that the MS severity score has better metric properties that correct the EDSS for disease duration ^39^.

Reports on association between lipid fractions levels and EDSS and MS severity are however inconsistent. Whereas some studies report that worsening EDSS and MS severity was associated with higher LDL-C and TGs but not HDL-C ^39, 40^, others showed the association between LDL-C and TG levels and EDSS diminished after accounting for confounding but remained significant for MS severity ^40^. Moreover, other studies found no significant association between lipid fractions and MS severity or EDSS ^41, 42^. Indeed, confounding and reverse causality in observational studies cannot be entirely ruled out. In the current study, MR approach was used, which limited the potential bias associated with the presence of confounders.

No evidence of association was found between variants in the gene regions that mimic the cholesterol-dependent and cholesterol-independent pathways and MS severity. To the best of our knowledge, the impact of statin treatment on disability progression measured by the MS severity score has not yet been studied. A handful of studies have explored the impact of statins on disability progression measured by the EDSS; however, the results were inconclusive. Whereas the phase 2 MS-STAT trial reports an association ^1^, others found that statin treatment had no effect on the EDSS score ^43, 44^. The possible explanation for this apparent contradiction is that the phase 2 MS-STAT trial had a larger sample size and the statin doses were larger than the doses in the latter two studies, indicating the possibility that higher doses of statins may effective to reduce the worsening of disability in patients with MS.

## limitations

This study has several limitations. First, the major lipid fractions (HDL-C, LDL-C and TG) are each heterogeneous groups of particles defined by differences in particle size, density, apoprotein content, migration characteristics and relationships to disease, and these subfractions differ in their risk profiles ^45^. This study was designed to investigate total blood lipid levels and thus did not consider whether there are subtypes of these fractions (e.g. LDL sub-particles) ^45, 46^ that might play different roles in MS risk or severity. Second The current study is unable to determine the underlying mechanism(s) for the potential causal relationship between RAC2 and MS risk; however, it is hoped that the findings presented may motivate further basic science investigations. Thirdly, we cannot exclude the possibility that the absence of a causal link between statins and MS severity is due to other pathways unrelated to Rho GTPases or *HMGCR* inhibition, which we could not investigate here because such pathways remain to be identified. Finally, although reverse causation MR was not performed to determine whether MS severity is causally associated with alterations in lipid levels, MR-Steiger results indicated that the assumption of causal directionality was accurate.

## Conclusion

Taken together, evidence from this study supports the existence of a causal effect of HDL-C on MS risk and shows that there is no reverse causation; however, no evidence for the causal role of LDL-C and TGs on MS risk was found. The MR findings suggest that *RAC2* is a potent genetic modifier of MS risk. Since it has been reported to mediate some of the pleiotropic effect of statins, we suggest that statins reduce MS risk via a *RAC2*-related mechanism (cholesterol-independent pathway). No evidence was found of a causal effect for lipid-related traits on MS severity. Finally, the current genetic evidence did not support the repurposing of statin treatment to reduce severity.

## Data Availability

The GWAS summary data used in this article are available at the URLs as follows: Lipid fractions (GLGC) http://csg.sph.umich.edu/willer/public/lipids2013/ ; whole blood cis-eQTL (eQTLgen consortium) https://www.eqtlgen.org/cis-eqtls.html ; whole blood cis-seQTL (GTx consortium) https://www.gtexportal.org/home/datasets ;HDL-C (Neale Lab UK Biobank GWAS) https://docs.google.com/spreadsheets/d/1kvPoupSzsSFBNSztMzl04xMoSC3Kcx3CrjVf4yBmESU/edit#gid=178908679 ; MS risk (discovery) https://imsgc.net/?page_id=31 ; MS severity data is available upon request to the corresponding author of the original publication (Sawcer et al., 2011), Professor Jacob McCauley.

## Acknowledgements

We would like to sincerely thank the IMSGC, GLGC, GTEx and Neale Lab consortia for access to their summary statistics data. We also thank Professor Jacob McCauley and Doctor Ashley Beechamand for their assistance with the MS severity dataset.

## Data Availability Statement

The GWAS summary data used in this article are available at the URLs as follows: Lipid fractions (GLGC) http://csg.sph.umich.edu/willer/public/lipids2013/ ; whole blood cis-eQTL (eQTLgen consortium) https://www.eqtlgen.org/cis-eqtls.html ; whole blood cis-seQTL (GTEx consortium) https://www.gtexportal.org/home/datasets; HDL-C (Neale Lab UK Biobank GWAS) https://docs.google.com/spreadsheets/d/1kvPoupSzsSFBNSztMzl04xMoSC3Kcx3CrjVf4yBmESU/edit#gid=178908679 ; MS risk (discovery) https://imsgc.net/?page_id=31 ; MS severity data is available upon request the corresponding author of the original publication (Sawcer et al., 2011) ^21^, Prof. Jacob McCauley.

## Authors’ contributions

Study concept, design and statistical analysis: Mona M. Almramhi. Mona M. Almramhi and Sandesh Chopade had access to all the data in the study and takes responsibility for the integrity of the data. Interpretation of results: all authors. Critical revision of the manuscript for important intellectual content: Mona M. Almramhi, Nicholas W Wood, Chris Finan, Amand Schmidt, Catherine S Storm and Rachel Coneys. Study supervision: Nicholas W Wood.

## Funding

The author(s) disclosed receipt of the following financial support for the research: M.M.A. is funded by the Faculty of Applied Medical Sciences, King Abdulaziz University, Jeddah, Saudi Arabia.C.F received additional support from the National Institute for Health Research University College London Hospitals Biomedical Research Centre. C.S.S. is funded by Rosetrees Trust, John Black Charitable Foundation and the University College London MBPhD Programme. A.F.S is supported by BHF grant PG/18/503383, and acknowledges support by grant R01 LM010098 from the National Institutes of Health (United States). D.A.K. is supported by an MBPhD Award from the International Journal of Experimental Pathology. R.C. is funded by Eisai, on the Wolfson-Eisai Neurodegeneration University College London PhD programme. N.W.W. is a National Institute for Health Research senior investigator. N.W.W. receives support from the National Institute for Health Research University College London Hospitals Biomedical Research Centre.

### Patient consent for publication

Not applicable.

## References

1. Chataway J, Schuerer N, Alsanousi A, et al. Effect of high-dose simvastatin on brain atrophy and disability in secondary progressive multiple sclerosis (MS-STAT): a randomised, placebo-controlled, phase 2 trial. The Lancet. 2014;383(9936):2213–21.

2. Greenwood J, Steinman L, Zamvil SS. Statin therapy and autoimmune disease: from protein prenylation to immunomodulation. Nature Reviews Immunology. 2006;6(5):358.

3. Weber MS, Steinman L, Zamvil SS. Statins—treatment option for central nervous system autoimmune disease? Neurotherapeutics. 2007;4(4):693–700.

4. Takemoto M, Liao JK. Pleiotropic effects of 3-hydroxy-3-methylglutaryl coenzyme a reductase inhibitors. Arteriosclerosis, thrombosis, and vascular biology. 2001;21(11):1712–9.

5. Wang C-Y, Liu P-Y, Liao JK. Pleiotropic effects of statin therapy: molecular mechanisms and clinical results. Trends in molecular medicine. 2008;14(1):37–44.

6. Neuhaus O, Stüve O, Zarnvil SS, Hartung H-P. Are statins a treatment option for multiple sclerosis? The Lancet Neurology. 2004;3(6):369–71.

7. Rikitake Y, Liao JK. Rho GTPases, statins, and nitric oxide. Circulation research. 2005;97(12):1232–5.

8. Zhu Y, Casey PJ, Kumar AP, Pervaiz S. Deciphering the signaling networks underlying simvastatin-induced apoptosis in human cancer cells: evidence for non-canonical activation of RhoA and Rac1 GTPases. Cell death & disease. 2013;4(4):e568–e.

9. Cordle A, Koenigsknecht-Talboo J, Wilkinson B, Limpert A, Landreth G. Mechanisms of statin-mediated inhibition of small G-protein function. Journal of Biological Chemistry. 2005;280(40):34202–9.

10. Hodge RG, Ridley AJ. Regulating Rho GTPases and their regulators. Nature reviews Molecular cell biology. 2016;17(8):496.

11. Yang G, Schooling CM. Investigating genetically mimicked effects of statins via HMGCR inhibition on immune-related diseases in men and women using Mendelian randomization. Scientific reports. 2021;11(1):1–7.

12. Bennett DA, Holmes MV. Mendelian randomisation in cardiovascular research: an introduction for clinicians. Heart. 2017:heartjnl-2016-310605.

13. Williams DM, Finan C, Schmidt AF, Burgess S, Hingorani AD. Lipid lowering and Alzheimer disease risk: a mendelian randomization study. Annals of neurology. 2020;87(1):30–9.

14. Willer CJ, Schmidt EM, Sengupta S, et al. Discovery and refinement of loci associated with lipid levels. Nature genetics. 2013;45(11):1274.

15. Consortium*† IMSG, ANZgene, IIBDGC, WTCCC2. Multiple sclerosis genomic map implicates peripheral immune cells and microglia in susceptibility. Science. 2019;365(6460):eaav7188.

16. Hemani G, Tilling K, Davey Smith G. Correction: orienting the causal relationship between imprecisely measured traits using GWAS summary data. PLoS genetics. 2017;13(12):e1007149.

17. Cho Y, Haycock PC, Sanderson E, et al. Exploiting horizontal pleiotropy to search for causal pathways within a Mendelian randomization framework. Nature communications. 2020;11(1):1–13.

18. Almramhi MM, Storm CS, Kia DA, Coneys R, Chhatwal BK, Wood NW. The role of body fat in multiple sclerosis susceptibility and severity: A Mendelian randomisation study. Multiple Sclerosis Journal. 2022:13524585221092644.

19. Võsa U, Claringbould A, Westra H-J, et al. Large-scale cis-and trans-eQTL analyses identify thousands of genetic loci and polygenic scores that regulate blood gene expression. Nature genetics. 2021;53(9):1300–10.

20. Consortium G. The GTEx Consortium atlas of genetic regulatory effects across human tissues. Science. 2020;369(6509):1318–30.

21. Sawcer S, Hellenthal G, Pirinen M, et al. Genetic risk and a primary role for cell-mediated immune mechanisms in multiple sclerosis. Nature. 2011;476(7359):214.

22. Bowden J, Del Greco M F, Minelli C, Davey Smith G, Sheehan NA, Thompson JR. Assessing the suitability of summary data for two-sample Mendelian randomization analyses using MR-Egger regression: the role of the I 2 statistic. International journal of epidemiology. 2016;45(6):1961–74.

23. Zheng J, Brion MJ, Kemp JP, et al. The Effect of Plasma Lipids and Lipid-Lowering Interventions on Bone Mineral Density: A Mendelian Randomization Study. Journal of Bone and Mineral Research. 2020;35(7):1224–35.

24. Burgess S, Dudbridge F, Thompson SG. Combining information on multiple instrumental variables in Mendelian randomization: comparison of allele score and summarized data methods. Statistics in medicine. 2016;35(11):1880–906.

25. Burgess S, Bowden J, Fall T, Ingelsson E, Thompson SG. Sensitivity analyses for robust causal inference from Mendelian randomization analyses with multiple genetic variants. Epidemiology (Cambridge, Mass). 2017;28(1):30.

26. Benjamini Y, Hochberg Y. Controlling the false discovery rate: a practical and powerful approach to multiple testing. Journal of the Royal statistical society: series B (Methodological). 1995;57(1):289–300.

27. Yuan S, Xiong Y, Larsson SC. An atlas on risk factors for multiple sclerosis: a Mendelian randomization study. Journal of Neurology. 2021;268(1):114–24.

28. Eshaghi A, Kievit RA, Prados F, et al. Applying causal models to explore the mechanism of action of simvastatin in progressive multiple sclerosis. Proceedings of the National Academy of Sciences. 2019;116(22):11020–7.

29. Tell RM, Kimura K, Palić D. Rac2 expression and its role in neutrophil functions of zebrafish (Danio rerio). Fish & shellfish immunology. 2012;33(5):1086–94.

30. Saoudi A, Kassem S, Dejean AS, Gaud G. Rho-GTPases as key regulators of T lymphocyte biology. Small GTPases. 2014;5(4):e983862.

31. Sironi M, Guerini FR, Agliardi C, et al. An evolutionary analysis of RAC2 identifies haplotypes associated with human autoimmune diseases. Molecular biology and evolution. 2011;28(12):3319–29.

32. Ramaswamy M, Dumont C, Cruz AC, et al. Cutting edge: Rac GTPases sensitize activated T cells to die via Fas. The Journal of Immunology. 2007;179(10):6384–8.

33. Fattouh R, Guo C-H, Lam GY, et al. Rac2-deficiency leads to exacerbated and protracted colitis in response to Citrobacter rodentium infection. PloS one. 2013;8(4):e61629.

34. Yang Q, Pan W, Qian L. Identification of the miRNA–mRNA regulatory network in multiple sclerosis. Neurological research. 2017;39(2):142–51.

35. Durant R, Klouche K, Delbosc S, et al. Superoxide anion overproduction in sepsis: effects of vitamin E and simvastatin. Shock. 2004;22(1):34–9.

36. Habib A, Shamseddeen I, Nasrallah MS, et al. Modulation of COX-2 expression by statins in human monocytic cells. The FASEB Journal. 2007;21(8):1665–74.

37. Baetta R, Lento S, Ghilardi S, et al. Atorvastatin reduces long pentraxin 3 expression in vascular cells by inhibiting protein geranylgeranylation. Vascular pharmacology. 2015;67:38–47.

38. Sawaya AP, Jozic I, Stone RC, et al. Mevastatin promotes healing by targeting caveolin-1 to restore EGFR signaling. JCI insight. 2019;4(23).

39. Weinstock-Guttman B, Zivadinov R, Mahfooz N, et al. Serum lipid profiles are associated with disability and MRI outcomes in multiple sclerosis. Journal of neuroinflammation. 2011;8(1):127.

40. Tettey P, Simpson Jr S, Taylor B, et al. An adverse lipid profile is associated with disability and progression in disability, in people with MS. Multiple Sclerosis Journal. 2014;20(13):1737–44.

41. Conway DS, Thompson NR, Cohen JA. Influence of hypertension, diabetes, hyperlipidemia, and obstructive lung disease on multiple sclerosis disease course. Multiple Sclerosis Journal. 2017;23(2):277–85.

42. Zhang T, Tremlett H, Zhu F, et al. Effects of physical comorbidities on disability progression in multiple sclerosis. Neurology. 2018;90(5):e419–e27.

43. Sena A, Pedrosa R, Morais MG. Therapeutic potential of lovastatin in multiple sclerosis. Journal of neurology. 2003:754–5.

44. Soldán MMP, Pittock SJ, Weigand SD, Yawn BP, Rodriguez M. Statin therapy and multiple sclerosis disability in a population-based cohort. Multiple Sclerosis Journal. 2012;18(3):358–63.

45. Rádiková Ž, Penesová A, Vlček M, et al. Lipoprotein profiling in early multiple sclerosis patients: effect of chronic inflammation? Lipids in health and disease. 2020;19(1):1–10.

46. Zhornitsky S, McKay KA, Metz LM, Teunissen CE, Rangachari M. Cholesterol and markers of cholesterol turnover in multiple sclerosis: relationship with disease outcomes. Multiple sclerosis and related disorders. 2016;5:53–65.

