## Supplementary for "Exploring the Role of Plasma Lipids and Statins Interventions on Multiple Sclerosis Risk and Severity: A Mendelian Randomization Study"

#### MS severity data

The MS severity score is an algorithm that relates scores on the EDSS to the distribution of disability in patients with comparable disease durations [1]. The MS severity score is designed to predict disease severity.

We obtained the summary statistics data for MS severity, from Professor Jacob McCauley, the corresponding author of the original publication [2]. The MS severity data has been generated from a genome-wide scan performed in MS cases (7,069 cases) to identify genetic variants that might influence MS severity [2]. In this severity-based analysis, the MS cases obtained from the discovery phase of the primary analysis of susceptibility of the 2011 GWAS, which included 9,772 cases and 17,376 controls [2]. Of the 9,772 cases, the disease severity (as measured by MS severity score) was available for only 7,069 cases, thus association analyses were only performed on 7,069 cases [2]. No genetic variants with strong evidence (p-value $< 5 \times 10^{-8}$) for association with MS severity were identified in that severity-based analysis [2].

**Table S1:** The Rho GTPase family and cholesterol biosynthesis pathway (mevalonate pathway) gene list (*part 1*)

| HGNC symbol | Ensemble Gene ID | Description | Participant of | Genes - MS risk analyses | Genes - MS severity analyses |
|---|---|---|---|---|---|
| *PPAPDC2* | ENSG00000205808 | phosphatidic acid phosphatase type 2 domain containing 2 | Cholesterol biosynthesis | Yes | Yes |
| *HMGCS1* | ENSG00000112972 | 3-hydroxy-3-methylglutaryl-CoA synthase 1 (soluble) | Cholesterol biosynthesis | Yes | Yes |
| *TM7SF2* | ENSG00000149809 | transmembrane 7 superfamily member 2 | Cholesterol biosynthesis | Yes | Yes |
| *ARV1* | ENSG00000173409 | ARV1 homolog (S. cerevisiae) | Cholesterol biosynthesis | Yes | Yes |
| *IDI1* | ENSG00000067064 | isopentenyl-diphosphate delta isomerase 1 | Cholesterol biosynthesis | Yes | Yes |
| *HMGCR* | ENSG00000113161 | 3-hydroxy-3-methylglutaryl-CoA reductase | Cholesterol biosynthesis | Yes | Yes |
| *LBR* | ENSG00000143815 | lamin B receptor | Cholesterol biosynthesis | Yes | Yes |
| *PMVK* | ENSG00000163344 | phosphomevalonate kinase | Cholesterol biosynthesis | Yes | Yes |
| *FDFT1* | ENSG00000079459 | farnesyl-diphosphate farnesyltransferase 1 | Cholesterol biosynthesis | Yes | Yes |
| *DHCR24* | ENSG00000116133 | 24-dehydrocholesterol reductase | Cholesterol biosynthesis | Yes | Yes |
| *MVK* | ENSG00000110921 | mevalonate kinase | Cholesterol biosynthesis | Yes | No |
| *FDPS* | ENSG00000160752 | farnesyl diphosphate synthase | Cholesterol biosynthesis | Yes | Yes |
| *ACAT2* | ENSG00000120437 | acetyl-CoA acetyltransferase 2 | Cholesterol biosynthesis | Yes | Yes |
| *GGPS1* | ENSG00000152904 | geranylgeranyl diphosphate synthase 1 | Cholesterol biosynthesis | Yes | Yes |
| *CYP51A1* | ENSG00000001630 | cytochrome P450, family 51, subfamily A, polypeptide 1 | Cholesterol biosynthesis | Yes | Yes |
| *HSD17B7* | ENSG00000132196 | hydroxysteroid (17-beta) dehydrogenase 7 | Cholesterol biosynthesis | Yes | Yes |
| *MVD* | ENSG00000167508 | mevalonate (diphospho) decarboxylase | Cholesterol biosynthesis | Yes | Yes |
| *SQLE* | ENSG00000104549 | squalene epoxidase | Cholesterol biosynthesis | Yes | Yes |
| *SC5DL* | ENSG00000109929 | sterol-C5-desaturase | Cholesterol biosynthesis | Yes | Yes |
| *LSS* | ENSG00000160285 | lanosterol synthase (2,3-oxidosqualene-lanosterol cyclase) | Cholesterol biosynthesis | Yes | Yes |
| *MSMO1* | ENSG00000052802 | methylsterol monooxygenase 1 | Cholesterol biosynthesis | NO | NO |
| *EBP* | ENSG00000147155 | emopamil binding protein (sterol isomerase) | Cholesterol biosynthesis | NO | NO |
| *IDI2* | ENSG00000148377 | isopentenyl-diphosphate delta isomerase 2 | Cholesterol biosynthesis | NO | NO |
| *NSDHL* | ENSG00000147383 | NAD(P) dependent steroid dehydrogenase-like | Cholesterol biosynthesis | NO | NO |
| *DHCR7* | ENSG00000172893 | 7-dehydrocholesterol reductase | Cholesterol biosynthesis | Yes | NO |
| *RHOV* | ENSG00000104140 | ras homolog family member V | member of Rho GTPase | NO | NO |
| *RHOB** | ENSG00000143878 | ras homolog family member B | member of Rho GTPase | Yes | Yes |
| *CDC42** | ENSG00000070831 | cell division cycle 42 | member of Rho GTPase | Yes | Yes |
| *RHOG** | ENSG00000177105 | ras homolog family member G | member of Rho GTPase | Yes | Yes |

| | | | | | |
|---|---|---|---|---|---|
| *RHOD** | ENSG00000173156 | ras homolog family member D | member of Rho GTPase | Yes | NO |
| *RHOF** | ENSG00000139725 | ras homolog family member F (in filopodia) | member of Rho GTPase | Yes | Yes |
| *RHOQ** | ENSG00000119729 | ras homolog family member Q | member of Rho GTPase | Yes | Yes |
| *RAC2** | ENSG00000128340 | ras-related C3 botulinum toxin substrate 2 (rho family, small GTP binding protein Rac2) | member of Rho GTPase | Yes | Yes |
| *RAC1** | ENSG00000136238 | ras-related C3 botulinum toxin substrate 1 (rho family, small GTP binding protein Rac1) | member of Rho GTPase | Yes | Yes |
| *RHOH** | ENSG00000168421 | ras homolog family member H | member of Rho GTPase | Yes | Yes |
| *RHOBTB2* | ENSG00000008853 | Rho-related BTB domain containing 2 | member of Rho GTPase | Yes | Yes |

**Table S1:** The Rho GTPase family and cholesterol biosynthesis pathway (mevalonate pathway) gene list (*part 2*)

| HGNC symbol | Ensemble Gene ID | Description | Participant of | Genes - MS risk analyses | Genes - MS severity analyses |
|---|---|---|---|---|---|
| *RAC3** | ENSG00000169750 | ras-related C3 botulinum toxin substrate 3 (rho family, small GTP binding protein Rac3) | member of Rho GTPase | NO | NO |
| *RHOU* | ENSG00000116574 | ras homolog family member U | member of Rho GTPase | Yes | NO |
| *RHOBTB1* | ENSG00000072422 | Rho-related BTB domain containing 1 | member of Rho GTPase | Yes | Yes |
| *RND2** | ENSG00000108830 | Rho family GTPase 2 | member of Rho GTPase | NO | NO |
| *RHOC** | ENSG00000155366 | ras homolog family member C | member of Rho GTPase | Yes | Yes |
| *RND1** | ENSG00000172602 | Rho family GTPase 1 | member of Rho GTPase | Yes | Yes |
| *RND3** | ENSG00000115963 | Rho family GTPase 3 | member of Rho GTPase | NO | NO |
| *RHOA** | ENSG00000067560 | ras homolog family member A | member of Rho GTPase | NO | NO |
| *RHOJ** | ENSG00000126785 | ras homolog family member J | member of Rho GTPase | NO | NO |

The table includes 25 genes that were flagged as being involved in the cholesterol biosynthesis pathway in the Reactome database of human pathways and reactions (http://www.reactome.org). The 20 genes of the Rho GTPase family were extracted from Azzarelli, 2015 [3]. For each gene, the HGNC symbol (HUGO Gene Nomenclature Committee), Ensemble gene ID and description were derived from the Ensembles database (https://www.ensembl.org/index.html). In the last two columns, ‘Yes’ indicates that a gene was included in the MR analysis, while ‘NO’ indicates that a gene was not included in the MR analysis due to either no SNPs being robustly associated with the target gene at a p-value $< 5 \times 10^{-8}$ or the eQTL data being absent.

* Rho GTPases that undergo prenylation, a total of 16 family members, which are the primary focus of the current study.

**Table S 2:** MR analysis of the effect of lipid level on MS risk

| Lipid trait | Method | No. of SNPs | OR (95 % CI) | p-value | FDR | pleiotropy assessment | | | |
|---|---|---|---|---|---|---|---|---|---|
| | | | | | | Q p-value | $I^2$ (%) | MR-Egger intercept | MR-Egger intercept p-value |
| **HDL-C** | IVW | 118 | 1.144 (1.04,1.26) | 7.94E-03 | 2.38E-02 | | | | |
| **HDL-C** | MR Egger | 118 | 1.229 (1.02,1.48) | 3.03E-02 | | 6.38E-06 | 40.5 | -0.004 | 3.66E-01 |
| **HDL-C adjusted for LDL-C & TG** | MVMR | 118 | 1.255 (1.06,1.49) | 0.00913 | | | | | |
| **LDL-C** | IVW | 99 | 0.996 (0.9,1.1) | 9.36E-01 | 9.36E-01 | | | | |
| **LDL-C** | MR Egger | 99 | 1.002 (0.85,1.18) | 9.82E-01 | | 1.30E-09 | 52.5 | -5.00E-04 | 9.27E-01 |
| **LDL-C adjusted for HDL-C & TG** | MVMR | 99 | 1.024 (0.94,1.12) | 0.607 | | | | | |
| **TG** | IVW | 65 | 0.921 (0.81,1.04) | 2.00E-01 | 3.00E-01 | | | | |
| **TG** | MR Egger | 65 | 0.859 (0.7,1.05) | 1.36E-01 | | 1.50E-04 | 43.6 | 0.0046 | 3.70E-01 |
| **TG adjusted for HDL-C & LDL-C** | MVMR | 65 | 1.076 (0.88,1.32) | 0.472 | | | | | |

HDL-C, high-density lipoprotein cholesterol; LDL-C, low-density lipoprotein cholesterol; TG, triglyceride; No. of SNPs, the number of independent genome-wide significant single nucleotide polymorphisms; IVW, inverse-variance weighted; MVMR, multivariable MR; OR, odds ratio; CI, confidence interval; Q p-value, Cochran’s Q statistic; FDR, false discovery rate; $I^2$ (%) expresses the level of heterogeneity as a percentage.

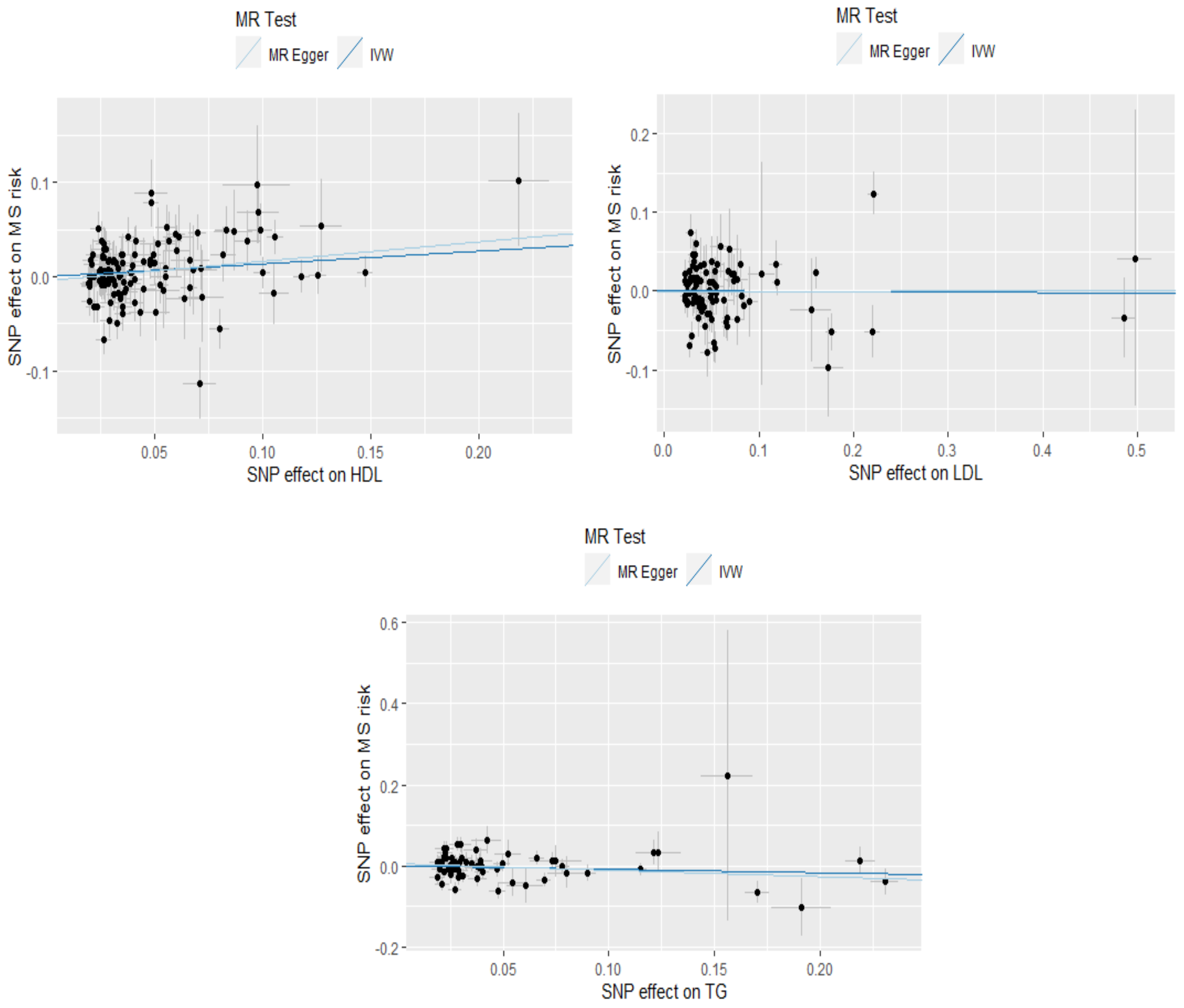

**Figure S 1:** Scatter plots for MR analyses showing the causal estimates of the lipid fractions on MS risk. The effect sizes of each genetic variant (with 95% confidence intervals) are represented by black points. The slope of each line shows the estimated MR effect for each method.

**Table S3:** MR analysis of the effect of lipid level on MS severity

| Trait | Method | No. of SNP | Beta (95 % CI) | p-value | FDR | Pleiotropy assessment | | | |
|---|---|---|---|---|---|---|---|---|---|
| | | | | | | Q p-value | I2 (%) | MR-Egger intercept | MR-Egger intercept p-value |
| **HDL-C** | IVW | 83 | -0.155 (-0.33,0.02) | 0.0847 | 0.254 | | | | |
| **HDL-C** | MR Egger | 83 | -0.039 (-0.49,0.41) | 0.867 | | 1 | 0 | -0.0067 | 0.546 |
| **HDL-C adjusted for LDL &TG** | MVMR | 83 | -0.059 (-0.4,0.28) | 0.735 | | | | | |
| **LDL-C** | IVW | 70 | -0.091 (-0.3,0.12) | 0.402 | 0.603 | | | | |
| **LDL-C** | MR Egger | 70 | -0.019 (-0.43,0.4) | 0.929 | | 0.805 | 0 | -0.005 | 0.684 |
| **LDL-C adjusted for HDL &TG** | MVMR | 70 | 0.012 (-0.35,0.37) | 9.49E-01 | | | | | |
| **TG** | IVW | 46 | -0.001 (-0.19,0.19) | 0.991 | 0.991 | | | | |
| **TG** | MR Egger | 46 | 0.08 (-0.39,0.54) | 0.738 | | 1 | 0 | -0.0055 | 0.661 |
| **TG adjusted for HDL &LDL** | MVMR | 46 | 0.035 (-0.22,0.29) | 7.88E-01 | | | | | |

The results reported as ORs with the 95% CI per 1-SD increase of lipid fractions. *For Abbreviations, see Table S2.*

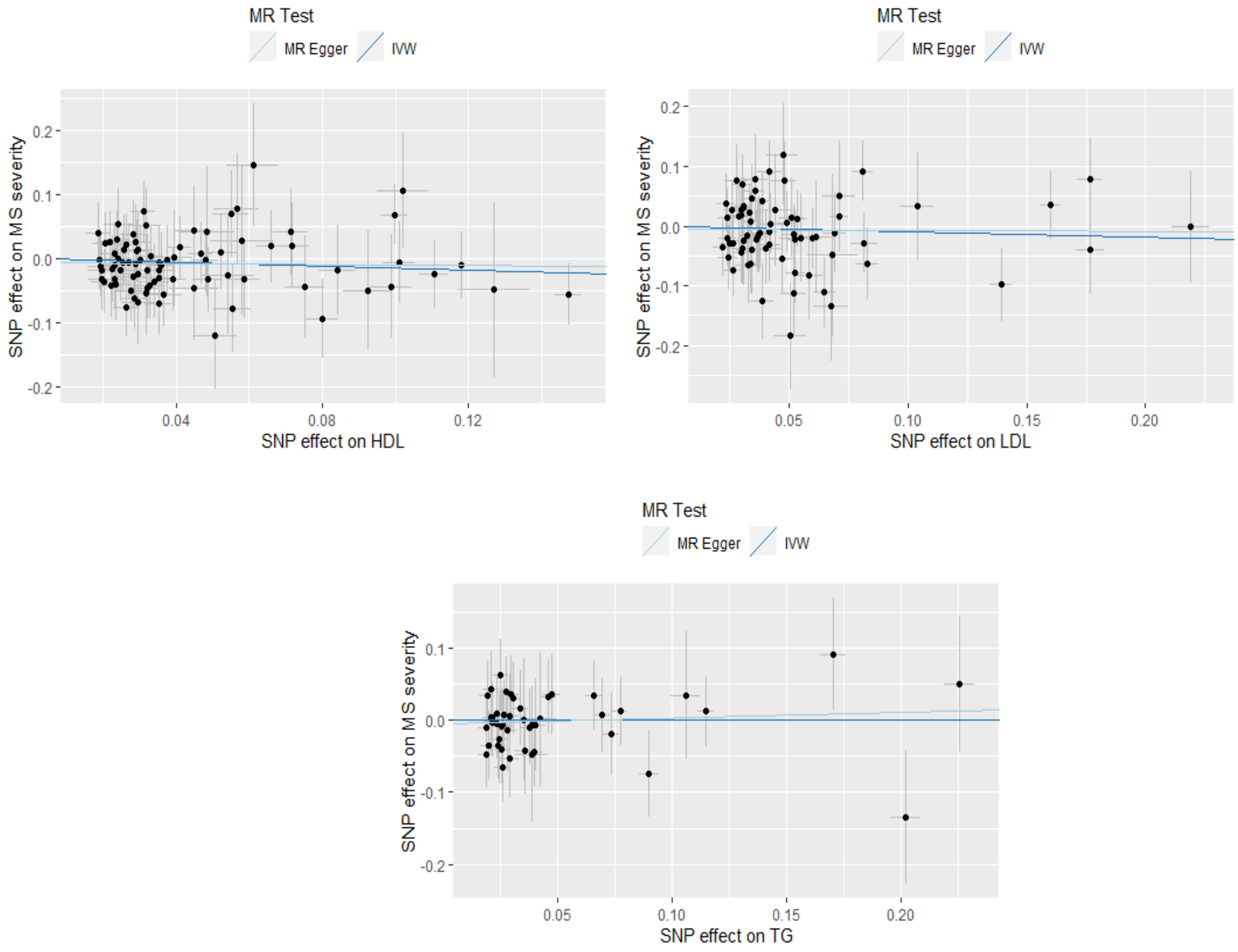


**Figure S2:** Scatter plots for MR analyses showing the causal estimates of the lipid fractions on MS severity. The effect sizes of each genetic variant (with 95% confidence intervals) are represented by black points. The slope of each line shows the estimated MR effect for each method.

**Table S4:** MR analysis of the effect of MS risk on lipid levels

| Outcome | Method | No. of SNP | Beta (95 % CI) | p-value | FDR | Pleiotropy assessment | | | |
|---|---|---|---|---|---|---|---|---|---|
| | | | | | | Q p-value | $I^2$ (%) | MR-Egger intercept | MR-Egger intercept p-value |
| **HDL-C** | IVW | 118 | -0.004 (-0.02,0.01) | 0.591 | 0.886 | | | | |
| **HDL-C** | MR Egger | 118 | -0.016 (-0.06,0.03) | 0.498 | | 4.05E-12 | 54 | 0.0012 | 0.593 |
| **LDL-C** | IVW | 118 | -0.008 (-0.02,0.01) | 0.295 | 0.885 | | | | |
| **LDL-C** | MR Egger | 118 | 0 (-0.04,0.05) | 0.991 | | 2.61E-07 | 44.5 | -0.0008 | 0.717 |
| **TG** | IVW | 119 | 0 (-0.01,0.01) | 0.961 | 0.961 | | | | |
| **TG** | MR Egger | 119 | 0.017 (-0.02,0.06) | 0.405 | | 2.55E-06 | 41.6 | -0.0017 | 0.37 |

The associations between genetically predicted MS risk and the lipid fractions are presented as 1-SD with 95% CI per 1-unit-higher log-odds of MS risk. *For Abbreviations, see Table S2.*

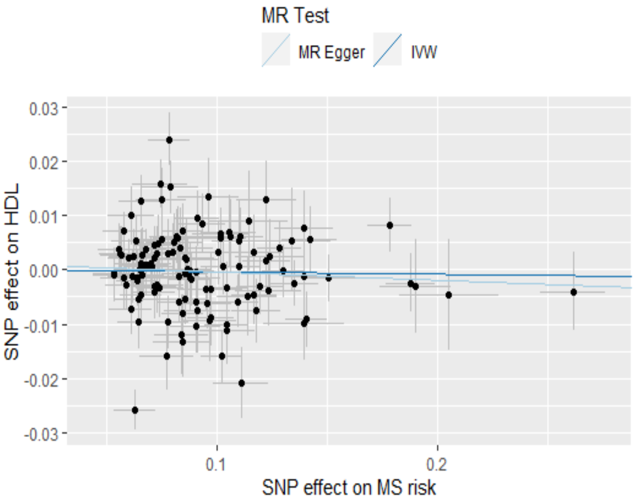
MR Test
MR Egger
IVW
SNP effect on HDL
0.03
0.02
0.01
0.00
-0.01
-0.02
-0.03
0.1
0.2
SNP effect on MS risk

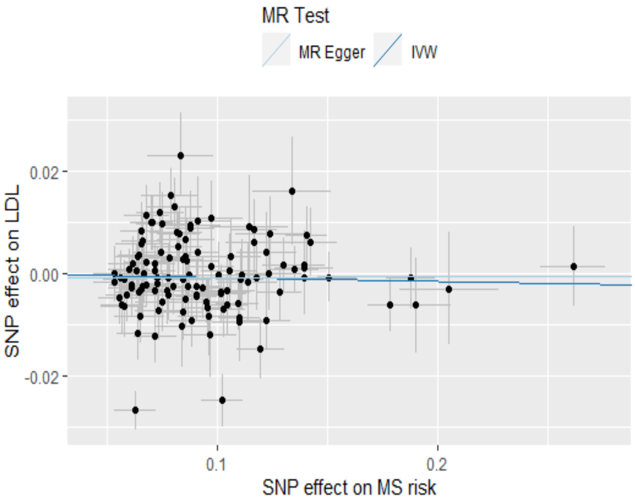
MR Test
MR Egger
IVW
SNP effect on LDL
0.02
0.00
-0.02
0.1
0.2
SNP effect on MS risk

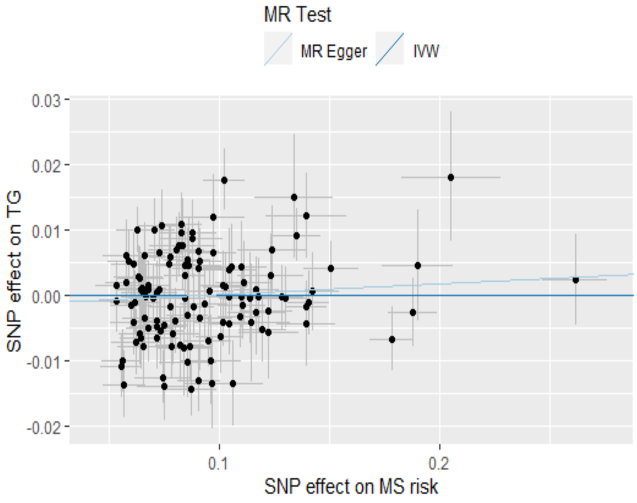
MR Test
MR Egger
IVW
SNP effect on TG
0.03
0.02
0.01
0.00
-0.01
-0.02
0.1
0.2
SNP effect on MS risk

**Figure S3:** Scatter plots for MR analyses showing the causal estimates of the MS risk on lipid fractions. The effect sizes of each genetic variant (with 95% confidence intervals) are represented by black points. The slope of each line shows the estimated MR effect for each method.

**Table S5**: MR estimates for the genetically mimicked effects of statins on MS risk (*part 1*)

| Trait | Method | No. of SNPs | OR (95 % CI) | p-value | FDR | pleiotropy assessment | | | |
|---|---|---|---|---|---|---|---|---|---|
| | | | | | | Q p-value | $I^2$ (%) | MR-Egger intercept | MR-Egger intercept p-value |
| ***ACAT2*** | IVW | 5 | 1.035(0.92,1.17) | 5.77E-01 | 8.74E-01 | | | | |
| ***ACAT2*** | MR Egger | 5 | 1.043(0.89,1.22) | 6.00E-01 | | 6.27E-01 | 0 | -0.0076 | 4.85E-01 |
| ***ARV1*** | IVW | 10 | 0.939(0.86,1.03) | 1.84E-01 | 7.38E-01 | | | | |
| ***ARV1*** | MR Egger | 10 | 0.941(0.84,1.05) | 2.96E-01 | | 6.79E-01 | 0 | -4.00E-04 | 9.52E-01 |
| ***CYP51A1*** | IVW | 3 | 1.029(0.89,1.19) | 6.99E-01 | 8.74E-01 | | | | |
| ***CYP51A1*** | MR Egger | 3 | 1.046(0.38,2.85) | 9.30E-01 | | 7.93E-01 | 0 | -0.0036 | 9.69E-01 |
| ***DHCR24*** | IVW | 9 | 1.053(0.92,1.2) | 4.37E-01 | 8.74E-01 | | | | |
| ***DHCR24*** | MR Egger | 9 | 1.024(0.73,1.44) | 8.90E-01 | | 9.61E-01 | 0 | 0.0072 | 8.12E-01 |
| ***DHCR7*** | Wald ratio | 1 | 0.431(0.18,1.02) | 5.68E-02 | 6.05E-01 | | | | |
| ***FDFT1*** | IVW | 25 | 0.984(0.95,1.02) | 4.07E-01 | 8.74E-01 | | | | |
| ***FDFT1*** | MR Egger | 25 | 0.979(0.94,1.02) | 2.62E-01 | | 3.33E-01 | 0.053 | 0.0058 | 6.12E-02 |
| ***FDPS*** | Wald ratio | 1 | 0.878(0.57,1.35) | 5.53E-01 | 8.74E-01 | | | | |
| ***GGPS1*** | IVW | 6 | 1.009(0.9,1.13) | 8.74E-01 | 8.74E-01 | | | | |
| ***GGPS1*** | MR Egger | 6 | 1.018(0.84,1.24) | 8.61E-01 | | 3.29E-01 | 0 | -0.0027 | 9.16E-01 |
| ***HMGCR*** | IVW | 3 | 1.178(0.89,1.56) | 2.50E-01 | 7.38E-01 | | | | |
| ***HMGCR*** | MR Egger | 3 | 1.074(0.09,12.56) | 9.55E-01 | | 3.33E-01 | 0 | 0.0101 | 9.41E-01 |
| ***HMGCS1*** | Wald ratio | 1 | 1.877(0.92,3.82) | 8.24E-02 | 6.05E-01 | | | | |
| ***HSD17B7*** | IVW | 9 | 1.061(0.83,1.36) | 6.40E-01 | 8.74E-01 | | | | |
| ***HSD17B7*** | MR Egger | 9 | 1.132(0.76,1.69) | 5.45E-01 | | 8.56E-02 | 0.36 | -0.0101 | 7.01E-01 |
| ***IDI1*** | IVW | 4 | 0.941(0.59,1.51) | 8.00E-01 | 8.74E-01 | | | | |
| ***IDI1*** | MR Egger | 4 | 0.493(0.21,1.17) | 1.10E-01 | | 4.97E-02 | 0.5 | 0.0491 | 1.13E-01 |
| ***LBR*** | IVW | 3 | 0.77(0.48,1.22) | 2.69E-01 | 7.38E-01 | | | | |
| ***LBR*** | MR Egger | 3 | 0.608(0,985.61) | 8.95E-01 | | 8.75E-01 | 0 | 0.0174 | 9.46E-01 |
| ***LSS*** | IVW | 15 | 0.993(0.94,1.05) | 8.03E-01 | 8.74E-01 | | | | |
| ***LSS*** | MR Egger | 15 | 1.006(0.93,1.09) | 8.82E-01 | | 3.67E-02 | 0.4 | -0.0028 | 6.84E-01 |

| | | | | | | | | | |
|---|---|---|---|---|---|---|---|---|---|
| ***MVD*** | IVW | 8 | 1.061(0.76,1.47) | 7.26E-01 | 8.74E-01 | | | | |
| ***MVD*** | MR Egger | 8 | 1.047(0.73,1.5) | 8.01E-01 | | 7.61E-03 | 0.6 | 0.0019 | 9.16E-01 |
| ***MVK*** | IVW | 3 | 1.079(0.78,1.49) | 6.39E-01 | 8.74E-01 | | | | |
| ***MVK*** | MR Egger | 3 | 1.237(0.14,10.68) | 8.47E-01 | | 7.63E-01 | 0 | -0.0148 | 8.87E-01 |

**Table S5**: MR estimates for the genetically mimicked effects of statins on MS risk (*part 2*)

| Trait | Method | No. of SNPs | OR (95 % CI) | p-value | FDR | pleiotropy assessment | | | |
|---|---|---|---|---|---|---|---|---|---|
| | | | | | | Q p-value | $I^2$ (%) | MR-Egger intercept | MR-Egger intercept p-value |
| ***PMVK*** | IVW | 4 | 0.877(0.69,1.11) | 2.81E-01 | 7.38E-01 | | | | |
| ***PMVK*** | MR Egger | 4 | 0.741(0.38,1.43) | 3.73E-01 | | 1.85E-01 | 0.11 | 0.0248 | 5.91E-01 |
| ***PPAPDC2*** | IVW | 9 | 1.065(0.97,1.17) | 2.02E-01 | 7.38E-01 | | | | |
| ***SC5DL*** | IVW | 4 | 0.989(0.89,1.1) | 8.43E-01 | 8.74E-01 | | | | |
| ***SC5DL*** | MR Egger | 4 | 0.993(0.82,1.2) | 9.40E-01 | | 3.21E-01 | 0 | -0.0013 | 9.62E-01 |
| ***SQLE*** | IVW | 5 | 1.024(0.82,1.28) | 8.38E-01 | 8.74E-01 | | | | |
| ***SQLE*** | MR Egger | 5 | 1.054(0.73,1.52) | 7.80E-01 | | 6.65E-01 | 0 | -0.0067 | 7.69E-01 |
| ***TM7SF2*** | IVW | 3 | 0.877(0.75,1.02) | 8.64E-02 | 6.05E-01 | | | | |
| ***TM7SF2*** | MR Egger | 3 | 1.068(0.37,3.06) | 9.03E-01 | | 8.01E-01 | 0 | -0.0489 | 6.56E-01 |
| ***PPAPDC2*** | MR Egger | 9 | 0.961(0.83,1.11) | 5.86E-01 | | 3.01E-01 | 0.045 | 0.0321 | 7.91E-02 |
| ***RAC1*** | IVW | 10 | 1.071(0.96,1.2) | 2.41E-01 | 5.41E-01 | | | | |
| ***RAC1*** | MR Egger | 10 | 1.218(0.9,1.66) | 2.08E-01 | | 7.11E-01 | 0 | -0.0168 | 3.56E-01 |
| ***RAC2*** | IVW | 15 | 0.861(0.78,0.95) | **3.80E-03** | **5.32E-02** | | | | |
| ***RAC2*** | MR Egger | 15 | 0.855(0.76,0.96) | **8.14E-03** | | 6.73E-01 | 0 | 0.0033 | 5.15E-01 |
| ***RHOB*** | IVW | 3 | 0.849(0.53,1.36) | 4.94E-01 | 5.83E-01 | | | | |
| ***RHOB*** | MR Egger | 3 | 1.053(0.07,15.89) | 9.70E-01 | | 6.53E-01 | 0 | -0.0172 | 8.66E-01 |
| ***RHOBTB1*** | IVW | 13 | 0.977(0.81,1.18) | 8.09E-01 | 8.09E-01 | | | | |
| ***RHOBTB1*** | MR Egger | 13 | 1.017(0.86,1.21) | 8.48E-01 | | 4.66E-02 | 0.4 | -0.0154 | 2.42E-01 |
| ***RHOBTB2*** | IVW | 3 | 0.713(0.51,1) | 4.94E-02 | 2.21E-01 | | | | |
| ***RHOBTB2*** | MR Egger | 3 | 0.868(0.36,2.11) | 7.56E-01 | | 5.70E-01 | 0 | -0.0273 | 5.61E-01 |
| ***RHOC*** | IVW | 5 | 1.038(0.83,1.3) | 7.44E-01 | 8.02E-01 | | | | |
| ***RHOC*** | MR Egger | 5 | 1.167(0.75,1.82) | 4.94E-01 | | 5.00E-02 | 0.49 | -0.0268 | 5.62E-01 |
| ***RHOD*** | IVW | 2 | 0.544(0.29,1.03) | 6.30E-02 | 2.21E-01 | | | | |
| ***RHOF*** | IVW | 6 | 1.134(0.93,1.39) | 2.18E-01 | 5.41E-01 | | | | |
| ***RHOF*** | MR Egger | 6 | 0.873(0.43,1.79) | 7.12E-01 | | 6.80E-01 | 0 | 0.0523 | 4.43E-01 |
| ***RHOG*** | IVW | 4 | 1.126(0.85,1.49) | 4.07E-01 | 5.69E-01 | | | | |
| ***RHOG*** | MR Egger | 4 | 1.228(0.84,1.8) | 2.91E-01 | | 2.38E-01 | 0 | -0.0153 | 4.97E-01 |
| ***RHOH*** | IVW | 5 | 0.518(0.29,0.94) | 3.01E-02 | 2.11E-01 | | | | |
| ***RHOH*** | MR Egger | 5 | 1.733(0.74,4.07) | 2.06E-01 | | 4.14E-01 | 0 | -0.0905 | **2.41E-03** |
| ***RHOQ*** | IVW | 9 | 1.07(0.88,1.3) | 4.99E-01 | 5.83E-01 | | | | |
| ***RHOQ*** | MR Egger | 9 | 0.909(0.66,1.25) | 5.56E-01 | | 5.79E-01 | 0 | 0.0261 | 1.70E-01 |

| | | | | | | | | | |
|---|---|---|---|---|---|---|---|---|---|
| ***RHOU*** | IVW | 3 | 1.161(0.89,1.51) | 2.70E-01 | 5.41E-01 | | | | |
| ***RHOU*** | MR Egger | 3 | 1.327(0.49,3.61) | 5.79E-01 | | 6.08E-01 | 0 | -0.0196 | 7.59E-01 |
| ***RND1*** | Wald ratio | 1 | 0.805(0.5,1.3) | 3.77E-01 | 5.69E-01 | | | | |
| ***CDC42*** | IVW | 10 | 1.054(0.93,1.19) | 3.98E-01 | 5.69E-01 | | | | |
| ***CDC42*** | MR Egger | 10 | 1.069(0.86,1.32) | 5.43E-01 | | 3.83E-02 | 0.45 | -0.0041 | 8.85E-01 |

Genes highlighted with orange encode proteins involved in cholesterol biosynthesis, while genes highlighted with green encode members of the Rho family. Pleiotropy assessment cannot be conducted for instruments consisting of ≤ 2 independent SNPs as it requires > 2 SNPs. *For Abbreviations, see Table S2.*

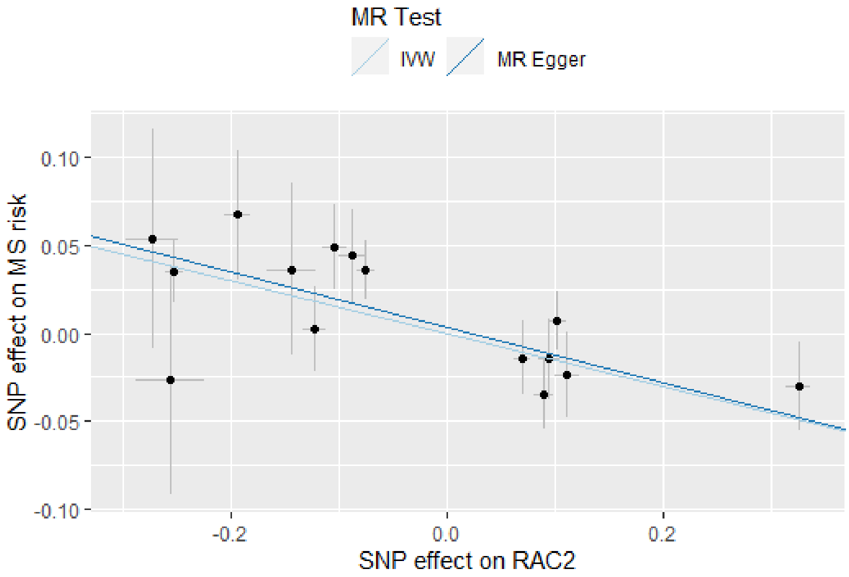


**Figure S4:** Scatter plots for MR analyses showing the causal estimates of *RAC2* on MS risk. The effect sizes of each genetic variant (with 95% confidence intervals) are represented by black points. The slope of each line shows the estimated MR effect for each method.

**Table S6**: MR estimates for the genetically mimicked effects of statins on MS severity (*part 1*)

| Trait | Method | No. of SNPs | Beta (95 % CI) | p-value | FDR | pleiotropy assessment | | | |
|---|---|---|---|---|---|---|---|---|---|
| | | | | | | Q p-value | $I^2$ (%) | MR-Egger intercept | MR-Egger intercept p-value |
| ***ACAT2*** | IVW | 3 | -0.145(-0.55,0.26) | 0.486 | 0.936 | | | | |
| ***ACAT2*** | MR Egger | 3 | -0.111(-0.58,0.36) | 0.642 | | 0.375 | 0 | -0.0129 | 0.46 |
| ***ARV1*** | IVW | 3 | 0.034(-0.24,0.31) | 0.808 | 0.951 | | | | |
| ***ARV1*** | MR Egger | 3 | 0.234(-0.46,0.93) | 0.509 | | 0.475 | 0 | -0.0656 | 0.498 |
| ***CYP51A1*** | Wald ratio | 1 | -0.155(-0.63,0.32) | 0.524 | 0.936 | | | | |
| ***DHCR24*** | IVW | 2 | 0.149(-0.24,0.54) | 0.456 | 0.936 | | | | |
| ***FDFT1*** | IVW | 8 | 0.163(0,0.33) | 0.0541 | 0.936 | | | | |
| ***FDFT1*** | MR Egger | 8 | 0.151(-0.04,0.34) | 0.117 | | 0.102 | 0.34 | 0.0042 | 0.834 |
| ***FDPS*** | Wald ratio | 1 | 0.152(-1.22,1.52) | 0.828 | 0.951 | | | | |
| ***GGPS1*** | IVW | 2 | 0.003(-0.32,0.33) | 0.988 | 0.988 | | | | |
| ***HMGCR*** | Wald ratio | 1 | 0.274(-0.65,1.2) | 0.562 | 0.936 | | | | |
| ***HMGCS1*** | Wald ratio | 1 | -1.142(-3.19,0.9) | 0.273 | 0.936 | | | | |
| ***HSD17B7*** | Wald ratio | 1 | 0.163(-1.21,1.54) | 0.817 | 0.951 | | | | |
| ***IDI1*** | IVW | 3 | -0.11(-0.94,0.72) | 0.795 | 0.951 | | | | |
| ***IDI1*** | MR Egger | 3 | 6.549(0.91,12.19) | 0.0229 | | 0.257 | 0 | -0.3974 | 0.0192 |
| ***LBR*** | IVW | 3 | -0.399(-1.74,0.94) | 0.558 | 0.936 | | | | |
| ***LBR*** | MR Egger | 3 | -0.129(-5.54,5.28) | 0.963 | | 0.517 | 0 | -0.0196 | 0.915 |
| ***LSS*** | IVW | 5 | -0.133(-0.28,0.01) | 0.0715 | 0.936 | | | | |
| ***LSS*** | MR Egger | 5 | -0.134(-0.29,0.02) | 0.0942 | | 0.393 | 0 | 0.0007 | 0.98 |
| ***MVD*** | Wald ratio | 1 | 0.227(-0.94,1.39) | 0.702 | 0.947 | | | | |
| ***PMVK*** | Wald ratio | 1 | 0.482(-0.6,1.56) | 0.381 | 0.936 | | | | |
| ***PPAPDC2*** | IVW | 3 | 0.105(-0.14,0.35) | 0.404 | 0.936 | | | | |
| ***PPAPDC2*** | MR Egger | 3 | 0.212(-12.8,13.23) | 0.975 | | 0.969 | 0 | -0.0279 | 0.985 |
| ***SC5DL*** | IVW | 2 | 0.073(-0.26,0.4) | 0.665 | 0.947 | | | | |
| ***SQLE*** | Wald ratio | 1 | 0.038(-1.41,1.48) | 0.959 | 0.988 | | | | |
| ***TM7SF2*** | IVW | 2 | -0.135(-0.65,0.38) | 0.604 | 0.936 | | | | |
| ***CDC42*** | IVW | 2 | 0.012(-0.73,0.75) | 0.975 | 0.988 | | | | |
| ***RAC1*** | IVW | 3 | -0.398(-0.92,0.12) | 0.134 | 0.936 | | | | |
| ***RAC1*** | MR Egger | 3 | -0.765(-3.44,1.91) | 0.575 | | 0.545 | 0 | 0.0631 | 0.776 |
| ***RAC2*** | IVW | 5 | 0.095(-0.25,0.44) | 0.588 | 0.936 | | | | |

| | | | | | | | | | |
|---|---|---|---|---|---|---|---|---|---|
| ***RAC2*** | MR Egger | 5 | 0.099(-0.59,0.79) | 0.779 | | 0.79 | 0 | -0.0012 | 0.985 |
| ***RHOB*** | Wald ratio | 1 | -0.646(-2.79,1.5) | 0.555 | 0.936 | | | | |
| ***RHOBTB1*** | IVW | 5 | 0.347(-0.15,0.84) | 0.17 | 0.936 | | | | |
| ***RHOBTB1*** | MR Egger | 5 | 0.325(-0.65,1.3) | 0.512 | | 0.278 | 0 | 0.0042 | 0.956 |
| ***RHOBTB2*** | IVW | 2 | -0.467(-1.78,0.85) | 0.486 | 0.936 | | | | |
| ***RHOC*** | IVW | 2 | 0.211(-0.45,0.87) | 0.531 | 0.936 | | | | |
| ***RHOF*** | IVW | 2 | 0.113(-0.44,0.67) | 0.692 | 0.947 | | | | |

**Table S6**: MR estimates for the genetically mimicked effects of statins on MS severity (*part 2*)

| Trait | Method | No. of SNPs | Beta (95 % CI) | p-value | FDR | pleiotropy assessment | | | |
|---|---|---|---|---|---|---|---|---|---|
| | | | | | | Q p-value | $I^2$ (%) | MR-Egger intercept | MR-Egger intercept p-value |
| ***RHOG*** | IVW | 4 | -0.366 (-1.12,0.39) | 0.339 | 0.936 | | | | |
| ***RHOG*** | MR Egger | 4 | -0.231 (-2.1,1.63) | 0.809 | | 0.803 | 0 | -0.023 | 0.779 |
| ***RHOH*** | IVW | 2 | -0.85 (-2.25,0.55) | 0.235 | 0.936 | | | | |
| ***RHOQ*** | IVW | 4 | -0.317 (-0.99,0.36) | 0.358 | 0.936 | | | | |
| ***RHOQ*** | MR Egger | 4 | -0.27 (-1.68,1.14) | 0.708 | | 0.357 | 0 | -0.0054 | 0.94 |
| ***RND1*** | Wald ratio | 1 | 0.212 (-2.14,2.57) | 0.86 | 0.952 | | | | |

Genes highlighted with orange encode proteins involved in cholesterol biosynthesis, while genes highlighted with green encode members of the Rho family. Pleiotropy assessment cannot be conducted for instruments consisting of ≤ 2 independent SNPs as it requires > 2 SNPs. The results reported as log ORs with the 95% CI per 1-SD higher expression of the target gene in blood. *For Abbreviations, see Table S2.*

### References


1. Roxburgh R, Seaman S, Masterman T, et al. Multiple Sclerosis Severity Score: using disability and disease duration to rate disease severity. Neurology. 2005;64(7):1144-51.
2. Sawcer S, Hellenthal G, Pirinen M, et al. Genetic risk and a primary role for cell-mediated immune mechanisms in multiple sclerosis. Nature. 2011;476(7359):214.
3. Azzarelli R, Kerloch T, Pacary E. Regulation of cerebral cortex development by Rho GTPases: insights from in vivo studies. Frontiers in cellular neuroscience. 2015;8:445.
